# Prioritizing disease-causing metabolic genes by integrating metabolomics with whole exome sequencing data

**DOI:** 10.1101/2021.05.21.21257573

**Authors:** Michiel Bongaerts, Ramon Bonte, Serwet Demirdas, Hidde Huidekoper, Janneke Langendonk, Martina Wilke, Walter de Valk, Henk J. Blom, Marcel J.T. Reinders, George J. G. Ruijter

## Abstract

The integration of metabolomics data with sequencing data is a key step towards improving the diagnostic process for finding the disease-causing gene(s) in patients suspected of having an inborn error of metabolism (IEM). The measured metabolite levels could provide additional phenotypical evidence to elucidate the degree of pathogenicity for variants found in metabolic genes. We present a computational approach, called *Reafect*, that calculates for each reaction in a metabolic pathway a score indicating whether that reaction is being deficient or not. When calculating this score, *Reafect* takes multiple factors into account: the magnitude and sign of alterations in the metabolite levels, the reaction distances between metabolites and reactions in the pathway, and the biochemical directionality of the reactions. We applied *Reafect* to untargeted metabolomics data of 72 patient samples with a known IEM and found that in 80% of the cases the correct deficient enzyme was ranked within the top 5% of all considered enzyme deficiencies. Next, we integrated *Reafect* with *CADD* scores (a measure for variant deleteriousness) and ranked the potential disease-causing genes of 27 IEM patients. We observed that this integrated approach significantly improved the prioritization of the disease-causing genes when compared with the two approaches individually. For 15/27 IEM patients the correct disease-causing gene was ranked within the top 0.2% of the set of potential disease-causing genes. Together, our findings suggest that metabolomics data improves the identification of disease-causing genetic variants in patients suffering from IEM.

## Introduction

DNA sequencing methods such as whole exome sequencing (WES) and whole genome sequencing (WGS) are powerful techniques to identify the pathogenic genetic variant(s) in patients suspected of a genetic disease (Pronicka, et al., 2016) (Wright, et al., 2018) (Stavropoulos, et al., 2016). Nevertheless, a single WES typically generates tens of thousands of variants (Wright, et al., 2018). With the reduced costs for sequencing, WGS becomes increasingly popular, generating even a few million of variants per patient (Wright, et al., 2018). Numerous filtering strategies have been developed to reduce the number of variants which need to be manually inspected. The Combined Annotation Dependent Depletion (*CADD*) score is widely explored as one of these filtering strategies (Rentzsch, et al., 2018); prioritizing variants such as single nucleotide variants (SNV), deletions and insertions (InDels) in patients. *CADD* scores employ a machine learning based approach where 63 conservational - and functional genomic metrics are combined into a single metric. After various filtering steps, the investigator still needs to evaluate a substantial number of variants manually. The pathogenicity of these rare or novel variants is often unknown, leading to a clinically dissatisfactory classification.

Functional studies may provide evidence whether a variant of unknown significance should be considered pathogenic or not. For this purpose, metabolomics is catching more and more interest since it has the potential to resolve the degree of pathogenicity for genetic variants which are expected to have an effect on the patient’s metabolism, i.e. inborn errors of metabolism (IEM) (Kerkhofs, et al., 2020) (Alaimo, et al., 2020) (Linck, et al., 2020). Some strategies have already been developed for this purpose; Haijes et al. applied expert knowledge to develop an algorithm that matches metabolic signatures obtained from metabolomics with expected metabolic signatures caused by each IEM, thereby ranking potential enzymatic deficiencies (Haijes, et al., 2020). Similarly, Baumgartner et al. explored the use of classification algorithms to distinguish multiple IEM based on differences in metabolite levels (Baumgartner, et al., 2004). However, training such a classifier requires data from multiple patients having the same IEM and since more than a 1000 different IEM exist with an overall birth prevalence of 51 per 100.000 (Waters, et al., 2018) the creation of large cohorts is challenging, thereby hampering the use of classification algorithms. To overcome this limitation, Messa et al. explored the use of metabolic networks to simulate IEM specific metabolic profiles, which they then compared with real IEM profiles using a Siamese neural network to rank the most probable matching (simulated) IEM (Messa, et al., 2020). Another strategy involves the use of gene-metabolite sets for which an enrichment score can be calculated to rank potential affected genes (Kerkhofs, et al., 2020). Similarly, *MetPropagate* (Linck, et al., 2020) uses gene-metabolite set enrichment scores, but additionally propagates these scores through a protein-protein network to rank potentially affected genes. The main concern with these approaches is that enrichment scores require (Z-score) cutoffs for metabolite levels, potentially excluding subtle aberrations that do not exceed the thresholds. In a different approach, Pirhaji et al. developed a tool, called *PIUMet*, that integrates metabolomics data with other omics data (Pirhaji, et al., 2016). *PIUMet* automatically annotates mass spectrometry (MS) features while inferring disease-associated pathways using a prize-collecting Steiner Forest algorithm. Still, we believe that most approaches did not fully exploited some crucial interconnected characteristics of IEM, i.e.: 1) the direction (increased or decreased) and 2) the magnitude of alterations in the metabolite levels, 3) pathway information including the biochemical directionality of reactions, and 4) reaction distances between metabolites and reactions.

To integrate metabolomics in WES, and potentially WGS, analysis, we developed an algorithm, called *Reafect (***Rea**ction de**fect**). *Reafect* combines information of metabolic pathways from KEGG (Kanehisa, 2000) and the metabolite Z-scores obtained from annotated metabolomics data to calculate a *‘deficient reaction score’* for each reaction. Higher scores imply that there is more evidence of that reaction being deficient and vice versa. Our algorithm differs fundamentally from the approaches mentioned earlier, since it is solely based on pathway information, and uses the metabolite Z-scores in a continuous fashion without using cutoff values. *Reafect* furthermore takes the directionality of the reactions and the sign of the Z-scores into account when calculating the *deficient reaction scores*. We evaluated *Reafect*’s performance on 36 distinct IEM using 72 plasma samples from patients diagnosed with an IEM.

Since each reaction is associated with genes coding for the enzyme catalyzing that reaction, we used *Reafect*’s *deficient reaction scores* in combination with *CADD* scores as an integrated model for prioritizing potentially disease-causing metabolic genes. To evaluate this approach, we studied 27 IEM patients for which the pathogenic variant was identified and untargeted metabolomics data was obtained. This integrated model showed a significant improvement on ranking the correct disease-causing genes when compared with using solely *Reafect* or *CADD* scores.

## Results

### Reafect

An enzymatic deficiency generally leads to a build-up of the reaction substrate(s) and shortages of the product(s) formed by that reaction. Z-scores obtained from annotated metabolomics (see Methods) can be used to detect the accumulation of these substrates (i.e. positive Z-scores) as well as shortages of the products (i.e. negative Z-scores). Although the accumulation and shortage of metabolites occur for the metabolites directly involved in the deficient reaction, aberrant metabolite levels will also propagate through a biochemical pathway, leading to changes in metabolite levels that are multiple reaction steps away from the deficient reaction. We used this dogma to develop an algorithm, called *Reafect*, that calculates for each reaction in a pathway a score that reflects how deficient that reaction is. We called this score the *deficient reaction score* or S_R_ score (see Methods for details). To calculate this score, *Reafect* weighs metabolite levels (Z-scores) which are further away from the considered reaction to a lesser extent than metabolite levels closer to the putative reaction, since we assume that more distant metabolites give less information about the reaction deficiency. For this purpose, *Reafect* uses a weighted version of the observed Z-scores, called *‘effective Z-scores’*, and which are always relative to the considered reaction for which the *deficient reaction score* is calculated (see Figure 1). The *effective Z-score* is determined by calculating a total decay factor over a reaction path when going from the metabolite (with Z-score) to that reaction. The more steps away from the considered reaction, the more the observed Z-score is decayed, thereby resulting in a lower (absolute) *effective Z-score*. To constrain the number of model parameters, we used three different decay factors (*a,b,c*) and distinguished five different decay types: 1) a decay factor *a* for a metabolite with a positive Z-score taking a step downstream towards the considered reaction, 2) a decay factor *b* for a metabolite with a positive Z-score taking a step upstream towards the considered reaction, 3) a decay factor *a* for a metabolite with a negative Z-score taking a step upstream, 4) a decay factor *b* for a metabolite with a negative Z-score taking a step downstream and 5) a decay factor *c* for reversible reactions (independent of the Z-score sign) taking one step in the direction of the considered reaction. We want to emphasize that *Reafect* describes reaction paths as a chain of metabolite and reaction nodes (in a graph) to track all pathway information (see Figure 1). Consequently, a reaction step is either a step from metabolite to reaction, or from reaction to metabolite. For example, consider a metabolite with a positive Z-score which takes three downstream steps to get to the considered reaction (Figure 1a, *m*_*2*_ to *R*_*3*_). The *effective Z-score* for this metabolite would then be given by the Z-score multiplied by *a*^*3*^, thus having a total decay factor of *a*^*3*^. Similarly, if this metabolite had a negative Z-score the total decay factor for this reaction path would have been *b*^*3*^. Obviously, a reaction path could also be more complex, resulting for example in a total decay factor of *c*^*2*^ *b a*^*2*^. We justify the introduction of *a* and *b*, by realizing that when *a>b* the *effective Z-scores* remain relatively high for positive Z-scores located upstream of a deficiency, and the same holds for negative Z-scores downstream of the deficiency. The values of these decay factors (*a, b* and *c*) are selected using the metabolomics data from 72 IEM patient samples (see Section *Tuning the model parameters*). Subsequently, *Reafect* aggregates all *effective Z-scores* resulting in the *deficient reaction score* (or S_R_ score) where it takes into account whether a certain *effective Z-score* was located downstream or upstream of the considered reaction (see Methods, Equation 6).

**Figure 1.**
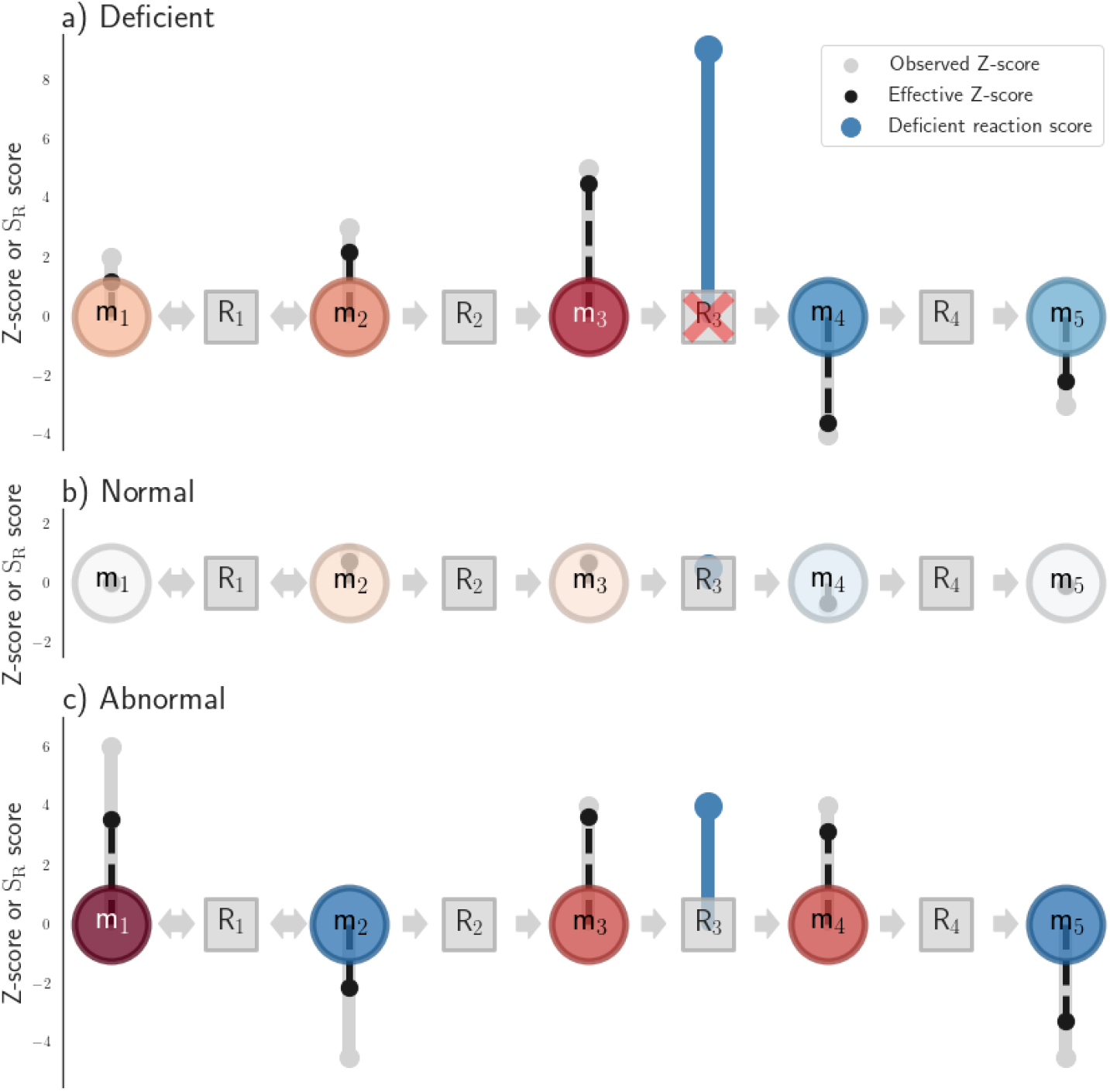
Illustration of *Reafect*. A circle indicates a metabolite and a square a reaction (node), with the horizontal arrows indicating the directionality of the reaction. The vertical grey bars (with dot) indicate the observed Z-scores; pointing upwards indicating a positive Z-score and vice versa. The black dotted bars indicate the *effective Z-score* from the perspective of reaction R_3_. Note that *Reafect* determines for each reaction a *deficient reaction score* but in these figures only the results are shown for R_3_. **A)** Reaction R_3_ is deficient. The *effective Z-scores* decay when going away from R_3_ as visualized by the reduced magnitude of the black bars. The *deficient reaction score*, illustrated by the blue bar on R_3_, is high since we observe net positive *effective Z-scores* upstream of R_3_ and net negative *effective Z-scores* downstream of R_3_. **B)** R_3_ is not deficient and metabolite Z-scores around the reaction are normal, thereby resulting in a low *deficient reaction score*. Note that the blue bar at R_3_ is small. **C)** R_3_ is not deficient, but has still a relatively high *deficient reaction score*. Note that although the observed Z-scores for m_3_ and m_4_ are equal, the resulting *effective Z-scores* are different since the decay of the Z-scores also depends on the biochemical directionality (and also applies to m_2_ and m_5_). Metabolite m_1_ has a relatively high observed Z-score, but its effective Z-scores is reduced since it is 5 reaction steps away from R_3._ *Reafect* calculates per side of the reaction the net *effective Z-scores*. For example, the *effective Z-scores* for m_4_ and m_5_ roughly counter balance each other when looking at the downstream side of R_3._ The upstream side has net positive *effective Z-scores*, therefore resulting in a positive *deficient reaction score*.

Finally, *Reafect* prioritizes all reactions by sorting the S_R_ scores on their magnitude, with higher scores indicating that a reaction is more likely to be deficient. Next to prioritizing the reactions, *Reafect* can prioritize enzymes and corresponding genes on their potential of being deficient. As enzymes can be involved in multiple reactions the final S_R_ score for an enzyme is taken to be the maximum S_R_ score of the set of reactions the enzyme may catalyze (Method).

### Tuning the model parameters

Per IEM patient, potential deficient enzymes were ranked by their maximum associated S_R_ score (Methods) and the rank of the true deficient enzyme in that patient was reported (Figure 4, *Absolute rank*). Since the total number of enzymes on which the ranking was based varied among the patients, we determined the percentile rank (PR) by dividing by the total number of enzymes multiplied by 100% (Methods). A lower PR indicates an improved ranking performance and vice versa. The overall performance of *Reafect* was measured by calculating how often a PR was smaller or equal than a predefined value across the 72 IEM patient samples. When increasing this predefined value a curve is generated as displayed in Figure 3. We used the area under this curve (AUC) to indicate the overall performance of *Reafect*, where higher AUCs imply better performances.

Since *Reafect* uses three model parameters (*a, b, c*), we used a parameter sweep over these parameters to explore how the performance (AUC) was affected. We performed a bootstrap procedure to obtain a robust performance AUC (Methods). Figure 2 shows these bootstrapped AUCs for each combination of (*a, b, c)*. For region *b > a, Reafect* performs less than for region *b < a*. This can be understood by realizing that when *a > b* the *effective Z-scores* for metabolites having positive Z-scores decay faster for downstream steps than for upstream steps (and the opposite for negative Z-scores), resulting in reduced evidence for the deficient reaction. Furthermore, for region *c < 0*.*5, Reafect’s* overall performance is poor. The highest performance was reached for *a* = 0.85, *b* = 0.35, and *c* = 0.75 (see Figure 2B). In further evaluations of *Reafect*, we set the parameters *a, b, c* to these values.

**Figure 2.**
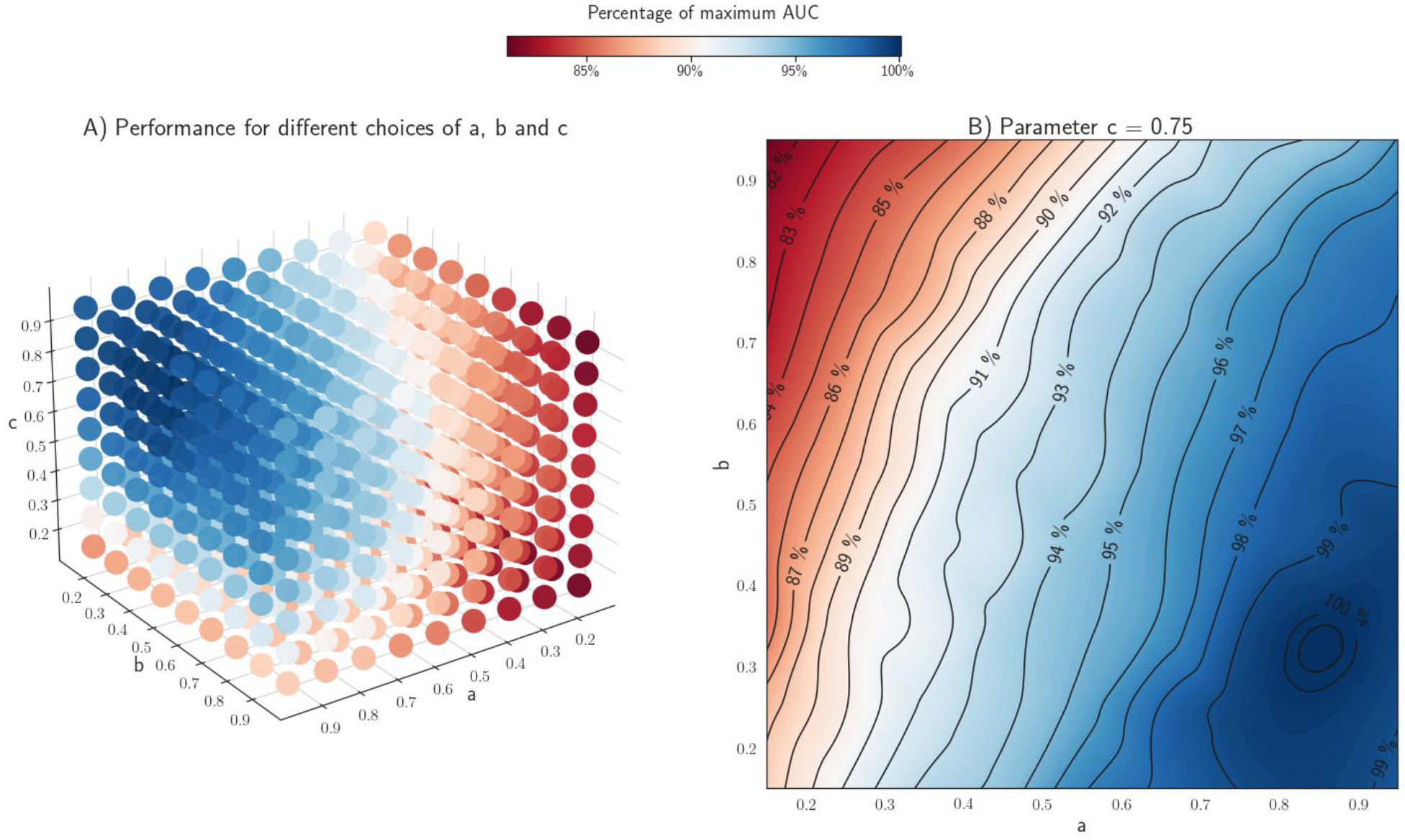
**A)** Bootstrapped AUCs (Methods) for different combinations of *Reafect*’s hyper parameters (*a,b,c*). The colors indicate the percentage of the maximum obtained AUC. **B)** Contour plot of the (cubic interpolated) bootstrapped AUCs while fixing *c=0*.*75* and varying *a* and *b*. The contour levels indicate the percentage of the maximum AUC reached at *a = 0*.*85, b = 0*.*35, c = 0*.*75*.

**Figure 3.**
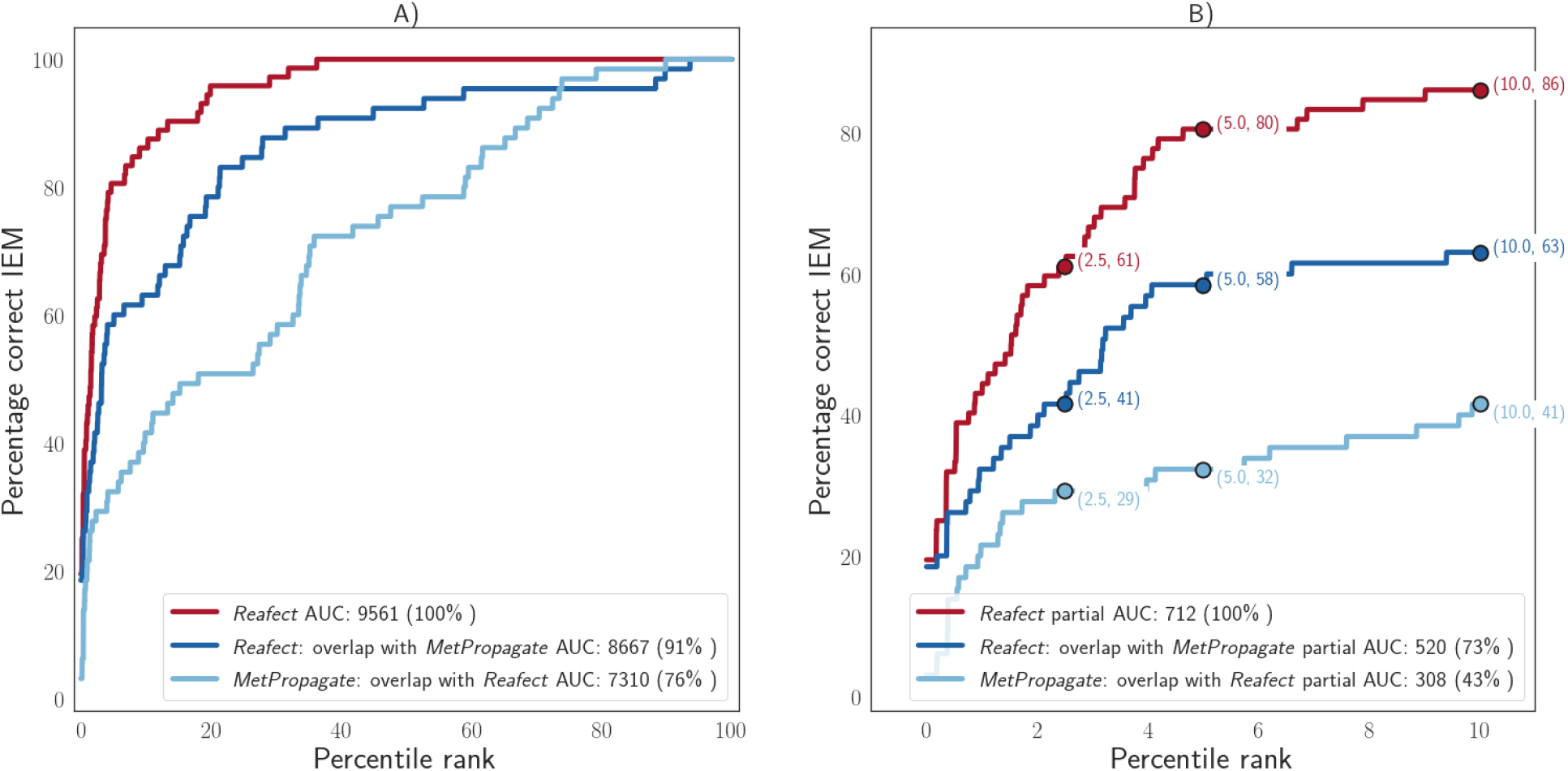
IEM ranking performances for different approaches as indicated by the legend. Each curve shows the percentage of IEM patient samples for which the percentile rank (PR) of the true enzyme deficiency is within the top *x* % (horizontal axis) of all considered enzyme deficiencies. Model settings for *Reafect*: a = 0.85, b = 0.35 and c = 0.75. **A)** Full performance curves. **B)** Performance curves with PR <= 10%. To perform a meaningful comparison between *Reafect* and *MetPropagate* a subset of the data was analyzed that contained only metabolites and genes that were included in both approaches (Methods). Note that this selection reduced the performance of *Reafect* to 91% of its original performance.

**Figure 4.**
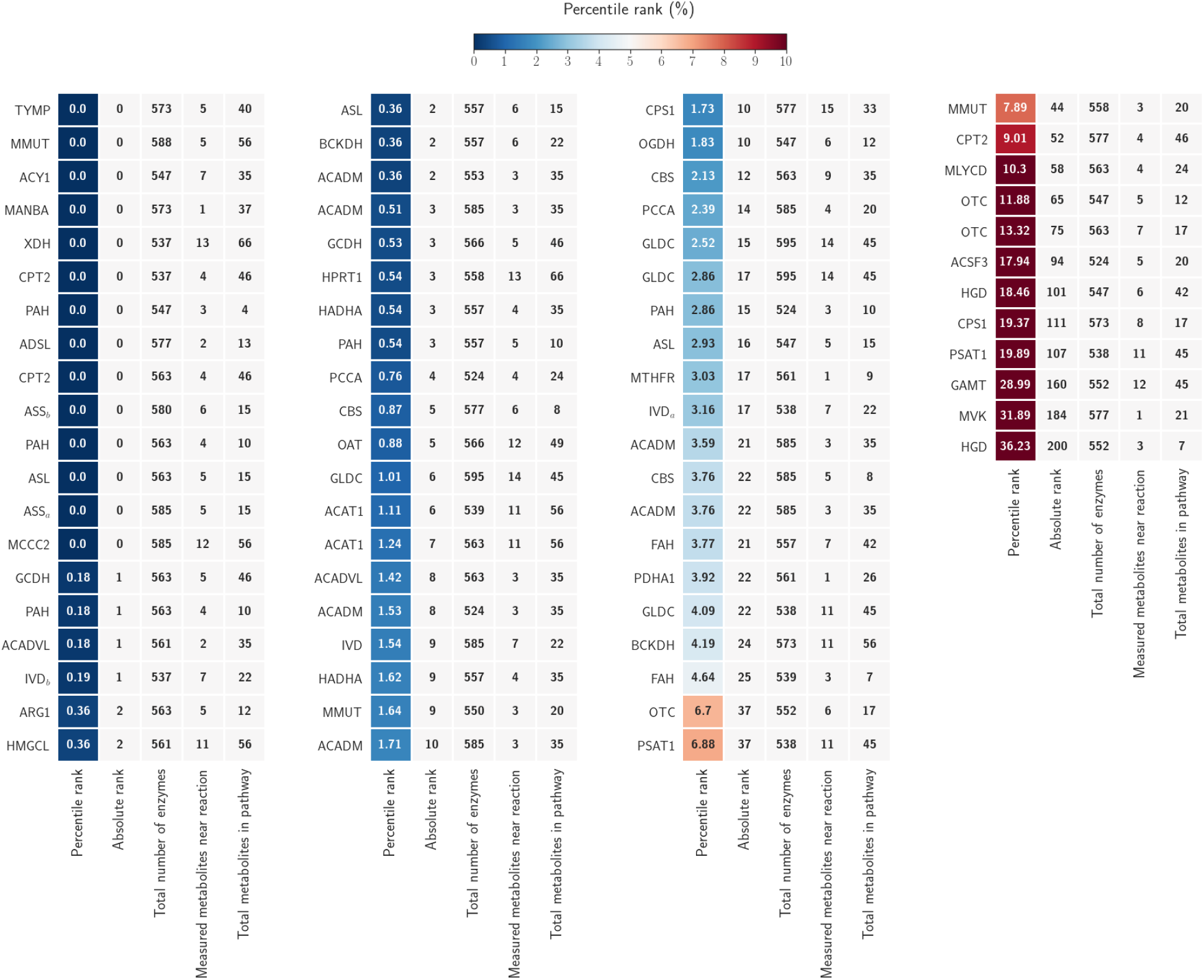
Detailed overview of the ranks of the correct IEM per patient. The first column indicates the PR (for the known deficient enzyme) for a given patient. Blue colors indicate PRs lower than 5%, orange/red colors indicate PRs above 5% (see color bar). The second column shows the absolute rank of the deficient enzyme. The third column indicates the total number of the ranks/ unique enzymes on which the ranking was based (this number varies across patients due to differences in metabolite annotations). The fourth column indicates the number of annotated metabolites in the pathway on which the *deficient reaction score* was based. The fifth column shows the total number of metabolites present in that pathway. For the HADHA gene, which encodes two enzymatic functions, we selected enzyme EC 1.1.1.211. The patient samples ASS_a_ and ASS_b_ originate from the same patient, but were acquired on different dates. The same holds for the samples IVD_a_ and IVD_b_.

### Enzyme ranking for IEM patients

We applied *Reafect* to 72 IEM patient samples and determined the percentile rank (PR) of the true enzyme deficiency. For 61% of these samples the PR was within the top 2.5% of all considered enzyme deficiencies, and for 80% of the samples the PR was within the top 5% (Figure 3). Additionally, we compared *Reafect* with *MetPropagate* (Linck, et al., 2020), while taking several factors into account such as overlapping metabolites and genes between the two approaches to objectively compare the performances (Methods). Based on 65 IEM patient samples, we found that *Reafect* has a 19% increase in the AUC when compared to *MetPropagate*. Considering that lower percentile ranks (<10%) are more interesting (Figure 3B), we observe that for this region the partial AUC of *Reafect* is 69% higher than the partial AUC of *MetPropagate*. A detailed overview of the PRs per IEM patient for both approaches can be found in Supplement 1.

Figure 4. shows a detailed overview of the results per IEM patient. From this figure it is clear that for the same IEM but different patients, *Reafect* can return different PRs. For example, one patient with maple syrup urine disease (BCKDH) has a PR of 0.36%, whereas for the other patient this is 4.19%. This can be explained by the difference in the magnitude of the Z-scores for the disease-related metabolites leucine and isoleucine, namely for the patient with the low rank Z = 6.7 and Z = 5.4, respectively, and for the patient with the higher rank these Z-scores were less extreme, Z = 2.49 and Z = 2.65, respectively. Similarly, for two patients having long-chain-3-hydroxyacyl-CoA dehydrogenase deficiency (HADHA), one has a PR of 0.54% and for the other this is 1.62%. Again, this difference in PRs can be understood by differences in for example 3-hydroxyhexadecanoylcarnitine, which had a Z-score of Z = 13.3 for the patient with the lower PR, while the other was more subtle with Z = 6.4. Also, one patient with carbamoyl phosphate synthetase I deficiency (CPS1) ranked at 1.73%, had Z = 1.8 for L-glutamine, while the other patient (ranked at 19.37%) seemed to have a normal L-glutamine level (Z = 0.2), thereby explaining also the difference between these ranks.

Some IEM were poorly ranked due to the absence of clear aberrations in the metabolomics data. For both patients with alkaptonuria (homogentisate 1,2-dioxygenase deficiency, HGD), homogentisic acid was not increased in our analysis (Z = 0.4 and Z = 0.5), which clarifies why *Reafect* poorly ranked these patients. The patient with mevalonate kinase deficiency (MVK) was also ranked poorly, which was a consequence of two reasons: 1) only one metabolite involved in calculating the S_R_ score i.e. mevalonic acid, was annotated in the metabolomics data and 2) the Z-score of this metabolite was Z = 0.7.

*Reafect* ranked the patient with arginase I deficiency (ARG1) at 0.36%. This was considered to be a relatively good ranking, since 14 metabolites were found to have a Z-score above 2.1, while the disease related metabolites arginine and ornithine had Z=2.1 and Z= -2.4 respectively. From a naive perspective we would expect about 14 other enzyme deficiencies to have lower (better) rank than arginase I. However, this relatively good performance can be explained by the fact that arginase I catalyzes the conversion of arginine into ornithine (plus urea), and the substrate (arginine) is increased while the product (ornithine) is reduced. Consequently, *Reafect* assigned a relatively high S_R_ score to this reaction. To strengthen this explanation, we used *Reafect* while flipping all Z-score signs (positive Z-scores become negative Z-scores and vice versa), and we observe that for this patient the PR increased from 0.36% to 26.11% (Supplement 2, Figure S3). This demonstrates that the obtained PR (0.36%) was a consequence of taking the Z-score signs and biochemical directionality into account.

Another interesting observation is the poor rank obtained for the patient having guanidinoacetate N-methyltransferase deficiency (GAMT). This patient was under treatment with creatine supplementation, which explains the poor rank. Although guanidinoacetate (Z = 3.1) was high in this patient, the presence of the high creatine level (Z = 6.7) led to high Z-scores on both sides of the GAMT reaction R01883 which reduces the S_R_ score, as can be observed in Equation 6 (Methods),

### Gene prioritization for IEM patients using *CADD* scores and *Reafect*

We hypothesized that potentially affected (metabolic) genes could be better prioritized when we combine the *CADD* (Phred) scores obtained from variants in WES data with the *deficient reaction scores* obtained from *Reafect*. Since an increase in both scores is expected to be associated with increased pathogenicity we chose to multiply the *deficient reaction score* with the maximum *CADD* score observed in the variants of the gene corresponding to that enzyme. Next, we used this combined score to rank the genes (Methods).

Since WES data was only available for two IEM patients, we evaluated this gene ranking based on two approaches: 1) using the WES background belonging to that patient if the WES was available (see asterisks in Table 1) and 2) using 15 random WES backgrounds while inserting the (known) disease-causing variant of the patient (Methods). Table 1 shows the PRs for 28 IEM patients for which the pathogenic variant was identified, using solely *Reafect*, solely *CADD* scores as well as the integrated approach. For 12/28 patients *Reafect* scored better than *CADD* (marked blue). For 21/28 and 20/28 patients, the integrated approach led to improved ranking when compared only to *Reafect* or *CADD* scores, respectively. Especially the gain in ranking performance for patients 5 (ACADVL), 7 (ACAT1), 15 and 16 (GLDC), and 23 (OGDH) is noteworthy (marked orange).

**Table 1.**
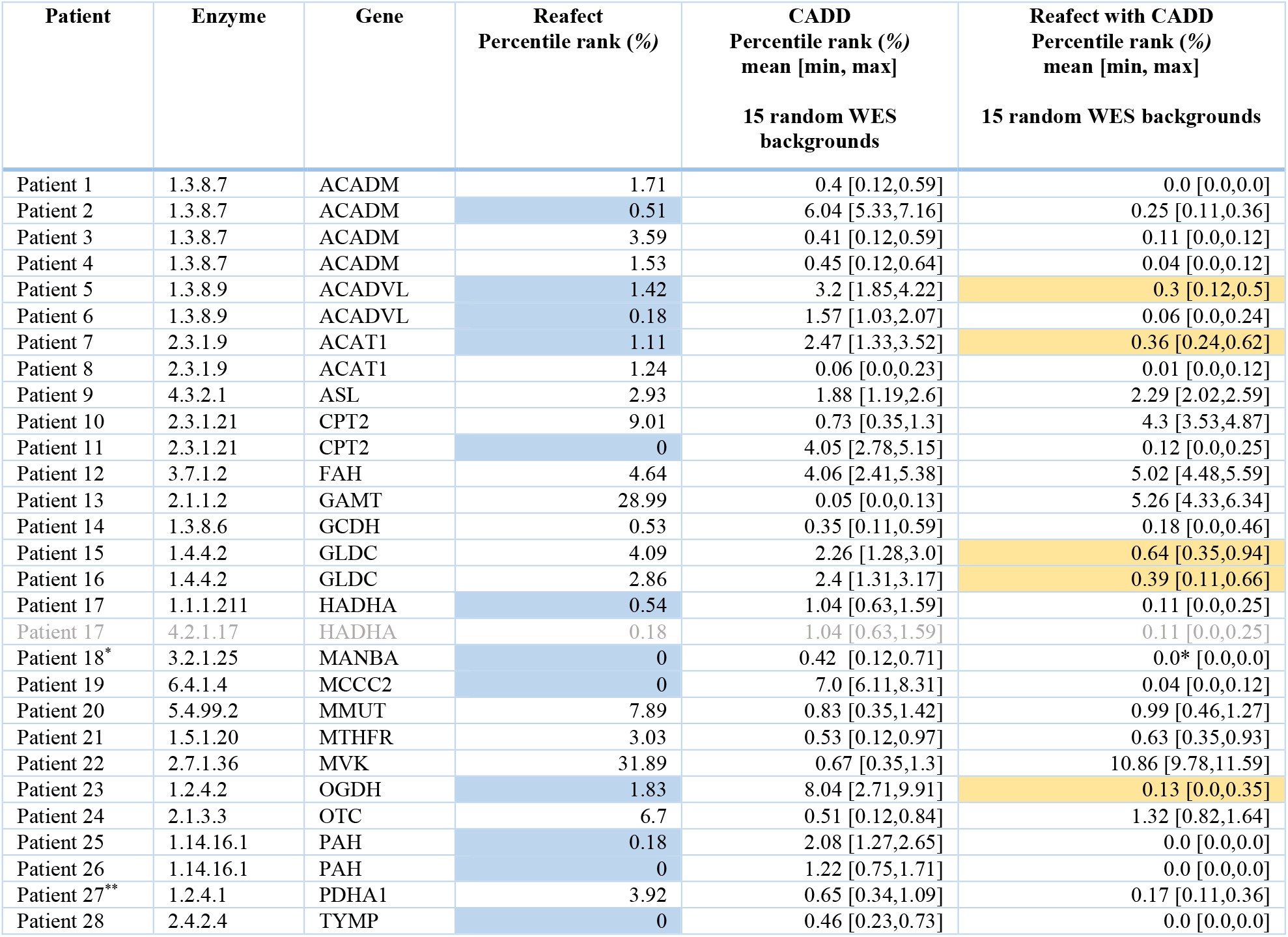
Overview of the IEM and disease-causing gene ranks for 28 IEM patients using *Reafect, CADD* scores and the integrated approach. The first column indicates the patient, second columns the deficient enzyme with EC identifier. The third columns refers to the affected gene. Next columns contain the PRs for each method as indicated by the column name; *Reafect* (only), *CADD* (only), and the integrated approach. The approaches using the 15 random WES backgrounds report the mean, minimum and maximum obtained PR across the 15 backgrounds. Blue marked results indicate that the PR of *Reafect* is lower than the PR of *CADD*. Orange marked results indicate a clear improvement of the integrated approach over the individual approaches. ^*^ The PR for *CADD* was 0.24% using the real WES, and 0.0% for *Reafect with CADD* using the real WES. ^**^ The PR for *CADD* was 0.58% using the real WES, and 0.35% for *Reafect with CADD* using the real WES.

| Patient | Enzyme | Gene | Reaffect<br>Percentile rank (%) | CADD<br>Percentile rank (%)<br>mean [min, max]<br>15 random WES<br>backgrounds | Reaffect with CADD<br>Percentile rank (%)<br>mean [min, max]<br>15 random WES backgrounds |
| --- | --- | --- | --- | --- | --- |
| Patient 1 | 1.3.8.7 | ACADM | 1.71 | 0.4 [0.12,0.59] | 0.0 [0.0,0.0] |
| Patient 2 | 1.3.8.7 | ACADM | 0.51 | 6.04 [5.33,7.16] | 0.25 [0.11,0.36] |
| Patient 3 | 1.3.8.7 | ACADM | 3.59 | 0.41 [0.12,0.59] | 0.11 [0.0,0.12] |
| Patient 4 | 1.3.8.7 | ACADM | 1.53 | 0.45 [0.12,0.64] | 0.04 [0.0,0.12] |
| Patient 5 | 1.3.8.9 | ACADVL | 1.42 | 3.2 [1.85,4.22] | 0.3 [0.12,0.5] |
| Patient 6 | 1.3.8.9 | ACADVL | 0.18 | 1.57 [1.03,2.07] | 0.06 [0.0,0.24] |
| Patient 7 | 2.3.1.9 | ACAT1 | 1.11 | 2.47 [1.33,3.52] | 0.36 [0.24,0.62] |
| Patient 8 | 2.3.1.9 | ACAT1 | 1.24 | 0.06 [0.0,0.23] | 0.01 [0.0,0.12] |
| Patient 9 | 4.3.2.1 | ASL | 2.93 | 1.88 [1.19,2.6] | 2.29 [2.02,2.59] |
| Patient 10 | 2.3.1.21 | CPT2 | 9.01 | 0.73 [0.35,1.3] | 4.3 [3.53,4.87] |
| Patient 11 | 2.3.1.21 | CPT2 | 0 | 4.05 [2.78,5.15] | 0.12 [0.0,0.25] |
| Patient 12 | 3.7.1.2 | FAH | 4.64 | 4.06 [2.41,5.38] | 5.02 [4.48,5.59] |
| Patient 13 | 2.1.1.2 | GAMT | 28.99 | 0.05 [0.0,0.13] | 5.26 [4.33,6.34] |
| Patient 14 | 1.3.8.6 | GCDH | 0.53 | 0.35 [0.11,0.59] | 0.18 [0.0,0.46] |
| Patient 15 | 1.4.4.2 | GLDC | 4.09 | 2.26 [1.28,3.0] | 0.64 [0.35,0.94] |
| Patient 16 | 1.4.4.2 | GLDC | 2.86 | 2.4 [1.31,3.17] | 0.39 [0.11,0.66] |
| Patient 17 | 1.1.1.211 | HADHA | 0.54 | 1.04 [0.63,1.59] | 0.11 [0.0,0.25] |
| Patient 17 | 4.2.1.17 | HADHA | 0.18 | 1.04 [0.63,1.59] | 0.11 [0.0,0.25] |
| Patient 18* | 3.2.1.25 | MANBA | 0 | 0.42 [0.12,0.71] | 0.0* [0.0,0.0] |
| Patient 19 | 6.4.1.4 | MCCC2 | 0 | 7.0 [6.11,8.31] | 0.04 [0.0,0.12] |
| Patient 20 | 5.4.99.2 | MMUT | 7.89 | 0.83 [0.35,1.42] | 0.99 [0.46,1.27] |
| Patient 21 | 1.5.1.20 | MTHFR | 3.03 | 0.53 [0.12,0.97] | 0.63 [0.35,0.93] |
| Patient 22 | 2.7.1.36 | MVK | 31.89 | 0.67 [0.35,1.3] | 10.86 [9.78,11.59] |
| Patient 23 | 1.2.4.2 | OGDH | 1.83 | 8.04 [2.71,9.91] | 0.13 [0.0,0.35] |
| Patient 24 | 2.1.3.3 | OTC | 6.7 | 0.51 [0.12,0.84] | 1.32 [0.82,1.64] |
| Patient 25 | 1.14.16.1 | PAH | 0.18 | 2.08 [1.27,2.65] | 0.0 [0.0,0.0] |
| Patient 26 | 1.14.16.1 | PAH | 0 | 1.22 [0.75,1.71] | 0.0 [0.0,0.0] |
| Patient 27** | 1.2.4.1 | PDHA1 | 3.92 | 0.65 [0.34,1.09] | 0.17 [0.11,0.36] |
| Patient 28 | 2.4.2.4 | TYMP | 0 | 0.46 [0.23,0.73] | 0.0 [0.0,0.0] |

To explore the overall differences in ranking performances between the three methods, we plotted the PRs in a boxplot (Figure 5). We removed the patient with guanidinoacetate N-methyltransferase deficiency from this analysis, arguing that the metabolic profile of this patient was not representative for this IEM because of the treatment. Using the Mann-Whitney U test, we observe that the performance between *Reafect* and *CADD* did not significantly differ (p-value > 0.05). However, the integrated approach significantly (Wilcoxon signed-rank test, p-value < 0.05) improved the ranking performance when compared with solely using *Reafect* or *CADD* scores. In other words, by combining the two scores we gained improved IEM ranking/ gene prioritization.

**Figure 5.**
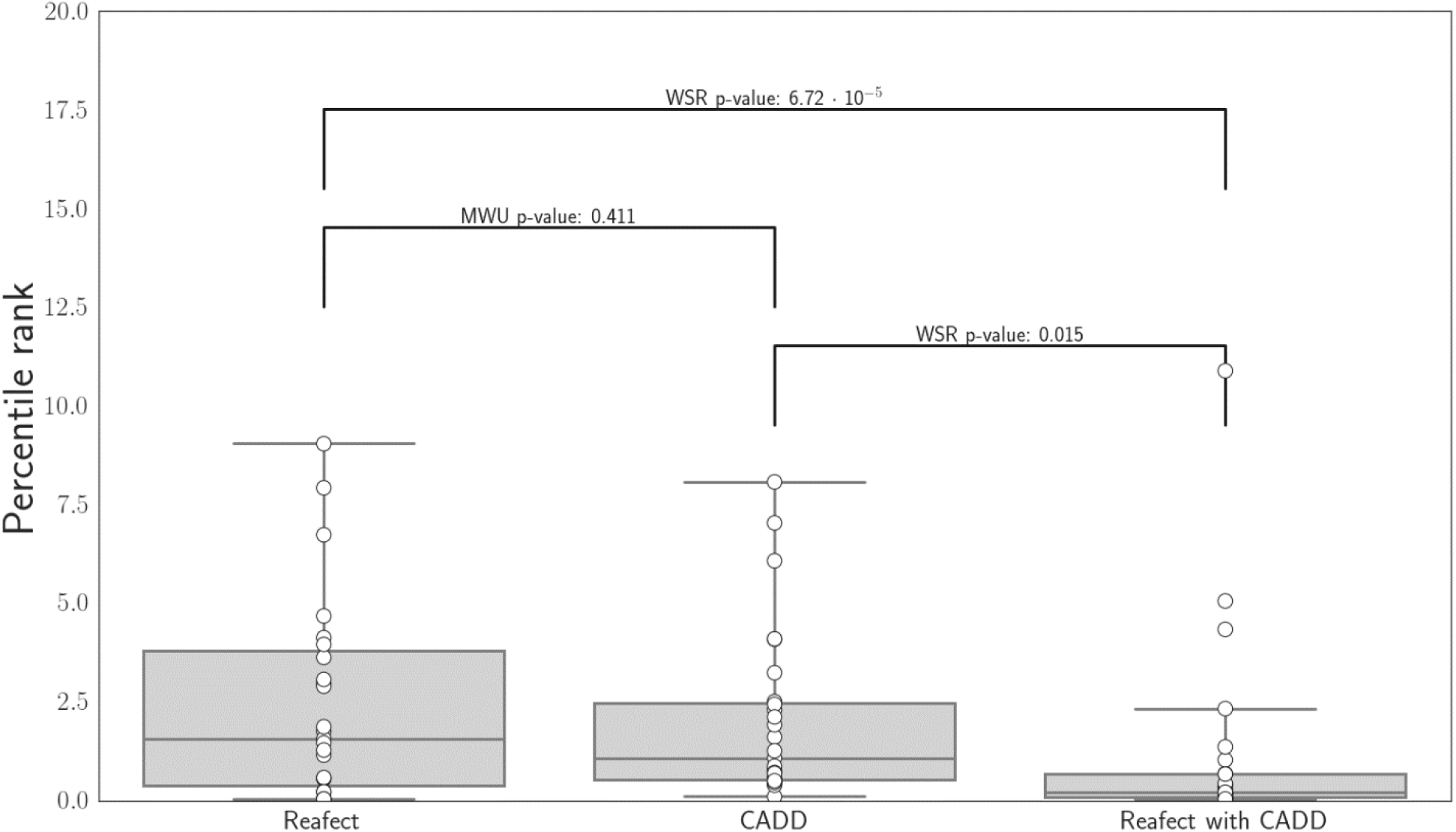
Boxplots of the percentile ranks (PRs) obtained from the different approaches; *Reafect* (only), *CADD* (only), and the integrated approach. For *CADD* and the integrated approach we used the average PR obtained from the 15 random WES. Significance was determined using the Wilcoxon signed-rank test (WSR) when comparing *Reafect with CADD* with *CADD* or *Reafect*. We used the Mann-Whitney U test (MWU) for comparing *CADD* with *Reafect*, arguing that the PRs for *CADD* and *Reafect* are independent since they are obtained from two separate datasets and approaches.

## Discussion

Our aim was to use metabolomics data as additional evidence for filtering genetic variants found in WES data. For this purpose, we developed *Reafect*, an algorithm that scores the efficacy of each reaction in a pathway. To calculate these scores, *Reafect* combines four types of information: 1) the magnitude and 2) sign of the metabolite Z-scores, 3) the biochemical directionality of reactions, and 4) the reaction distances between the metabolites and a reactions in a pathway. We observed that *Reafect* ranked the true deficient enzyme for 80% of the 72 IEM patient samples within the top 5% of all considered enzyme deficiencies. *Reafect* showed improved ranking performance when compared to *MetPropagate*. We anticipate that this improvement may at least partially be explained by three differences between *Reafect* and *MetPropagate*. First, since *MetPropagate* uses cutoff values for the metabolite Z-scores when calculating the enrichment scores, we expect relevant but subtle aberrant metabolites to be neglected. *Reafect* uses the Z-scores in a continuous fashion, therefore even subtle aberrations contribute to the *deficient reaction scores* and positively impact IEM ranking (see Supplement 6). Secondly, metabolite-gene set enrichment approaches only consider metabolites which have a direct relationship with a gene, such as well-known biomarkers. Metabolite levels which are multiple reaction steps away from the deficiency may still be informative but will not contribute to the enrichment score when these metabolites are not included in the metabolite-gene set. Thirdly, *MetPropagate*, and approaches like the ones suggested by Pirhaji et al. and Kerkhofs et al., do not explicitly take the directionality of reactions and the sign of metabolite levels (decreased/increased) into account. We showed that *Reafect*’s IEM ranking performance was greatly reduced by flipping the sign of the metabolite Z-scores (Supplement 2), emphasizing that the Z-score sign and reaction directionality are important for *Reafect*. We furthermore showed that by choosing optimal values for our model parameters (*a,b,c*), we were able to improve the IEM ranking performance. Since these model parameters relate to the interaction of the Z-score signs with reaction directionality, this again confirms that including this information is valuable.

Integration of metabolomics with WES was achieved by multiplying the maximum *deficient reaction scores* with the maximum *CADD* score found for each enzyme and corresponding gene respectively (Methods). This integrated approach resulted in a significant improvement of ranking the true disease-causing genes (see Figure 5), where the median percentile rank (PR) was 1.36% lower than the median PR obtained from *Reafect*, and was 0.87% lower than the median PR obtained from using solely *CADD* scores.

In reality the human metabolome is one interconnected network of metabolites and reactions. In this study we have chosen to use isolated metabolic modules/pathways for two reasons. First, the (KEGG) pathways are clusters of highly interdependent reactions, for which we expect multiple metabolite levels to be affected if a pathway contains an enzymatic deficiency. Secondly, the direct use of a complete metabolic network would introduce metabolic ‘hubs’ that would connect more distinct parts of the metabolism. This entanglement of pathways/reactions may have unwanted consequences for the *deficient reaction scores* since also less relevant metabolite Z-scores would be involved in the calculation of these scores. A negative consequence of using isolated modules/pathways might be that some important reactions are not included. Although the goal was to develop an algorithm with minimum manual adjustments, we needed to add several reactions, such as glycine conjugation and carnitine esterification, to increase the overlap between metabolites measured in plasma and the metabolites included in the pathways (Supplement 4).

*Reafect* also has some limitations. First, if not all metabolites in the KEGG pathway are measured and annotated, this may lead to wrong conclusions. A single metabolite with a relatively high Z-score will cause all (downstream) reactions to have high *deficient reaction scores*. The inclusion of more measured metabolites could prevent this behavior, since metabolite Z-scores with the same sign on both sides of the reaction reduce the *deficient reaction score* (Methods, Equation 6). The IEM ranking performance of *Reafect* is therefore affected by the number of metabolites being measured within each pathway. Secondly, *Reafect* is based on the assumption that IEM have the signature where substrates of the deficient reaction become more abundant and the products decrease in abundancy. In case such signature does not hold for a certain IEM, we expect *Reafect* to detect these kinds of IEM poorly. At last, *Reafect* ignores compartmentalization of different metabolic processes. A substantial number of metabolic reactions occur within certain compartments of the cell such as the mitochondrion. Similarly, different organs contain different sets of metabolic reactions, therefore the concentration of the affected metabolites for an IEM may be very different from the concentrations measured in plasma on which our Z-scores are based.

For most IEM patients with an identified disease-causing variant in this study, the putative gene was directly sequenced, and therefore no WES data was obtained. We inserted the identified disease-causing variant in 15 random WES backgrounds, to enable the inclusion of these patients in our study. We assumed that the average ranking obtained from these 15 backgrounds was still a good estimate of the ranking which would have been obtained when the real WES data was used. Due to our limited number of patients with real WES data (N=2), a reliable comparison between both rankings is not possible, and thus we cannot validate the accuracy of this assumption. Note that the PRs obtained from the real WES fall within the minimum and maximum PR obtained from the 15 WES backgrounds (Table 1).

*Reafect* uses only three decay factors (*a, b, c*) which we optimized using an overall performance metric (see Results, Figure 2). Ideally, these decay factors are optimized using a training set while using a separate validation set for evaluating the IEM ranking performances. Due to the low number of IEM patients included in this study we decided to use all samples for optimization and validation, arguing that splitting the dataset into a training - and validation set would lead to less accurate estimates of the decay factors and would give less insights into the overall performance of *Reafect* on distinct IEM. Note that we did use a bootstrap procedure to prevent overfitting of the decay factors (Methods). To further support our findings, we separately optimized the decay factors using a subset of the 72 IEM patient samples; the 44 samples for which the disease-causing variant was unknown. Using the same bootstrap procedure, we obtained an optimum close to the one found when using all 72 samples (Supplement 5).

We realize that the use of three decay factors is a simplification, and that these factors should ideally be reaction specific. Kinetic parameters, such as the Michaelis–Menten constant, could be used to establish such reaction dependent decay factor. Currently accurate kinetic parameters are only available for a subset of reactions. Besides the additional complexity introduced by these reaction specific decay factors, the use of just three decay factors offered us the opportunity to demonstrate the overall importance of choosing different decay factors for reaction directionality and the sign of the Z-score, as we clearly observed in Figure 2. Still, we anticipate that *Reafect*’s performance on ranking IEM/genes could improve when reaction specific decay factors are incorporated.

*Reafect* may not only be useful in the context of IEM but could be applicable in a wider context since the *deficient reaction scores* are a direct readout of potential accumulations and/or reductions of metabolites before/after a reaction. For example, *Reafect* is potentially useful in drug screening research for generating an overview of drug candidates which have the potential to inhibit metabolic enzymes. Namely, we expect that the inhibition of an enzyme by a drug will result in metabolic signatures similar to the ones caused by an IEM where the same enzyme is affected.

In conclusion, the integration of metabolomics data with WES data by using *Reafect’s deficient reaction scores* and *CADD* scores, significantly improved the prioritization of pathogenic genes in patients suffering from an IEM.

## Method

### Untargeted metabolomics data and Z-scores

Metabolomics data was obtained as described by Bonte et al. Samples obtained from IEM patients were measured in 20 separate batches and features were annotated using an in-house database having MS/MS spectra and retention times of each metabolite (Bonte, et al., 2019). For the 72 patient samples, a median of 119 annotated metabolites was obtained (when combining positive – and negative ion mode), and a minimum of 95 annotated metabolites was available for each sample. In agreement with national legislation and institutional guidelines, all patients or their guardians approved the possible anonymous use of the remainder of their samples for method validation and research purposes. The study was conducted in accordance with the Declaration of Helsinki. Z-scores were calculated using two different approaches: 1) metabolites which were annotated in at least 7 batches were merged, a Box-Cox transform was applied, normalized using *Metchalizer* (Bongaerts, et al., 2020) and the Z-scores were determined using a regression model with age and sex as covariates (Bongaerts, et al., 2020), 2) for metabolites which were annotated in less than 7 batches, the Z-scores were determined from 15 within-batch samples, where abundancies were first Box-Cox transformed and normalized using Probabilistic Quotient Normalization (PQN). When a metabolite was annotated in both positive- and negative ion mode, the Z-score of the ion mode with the largest median abundancy (over all samples) was taken. Since three technical replicates were measured for all patient samples, we used the average of these three Z-scores as the final Z-score (which was then transformed using Equation 1).

### Z-score transformation

To prevent extreme Z-scores to dominate the *deficient reaction scores*, we transformed the Z-score by applying:

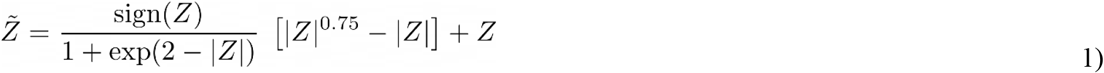

This transform behaves linear for the region 0 < |Z| < 2, but scales down Z-scores when |Z| >> 2 (Figure 6).

**Figure 6.**
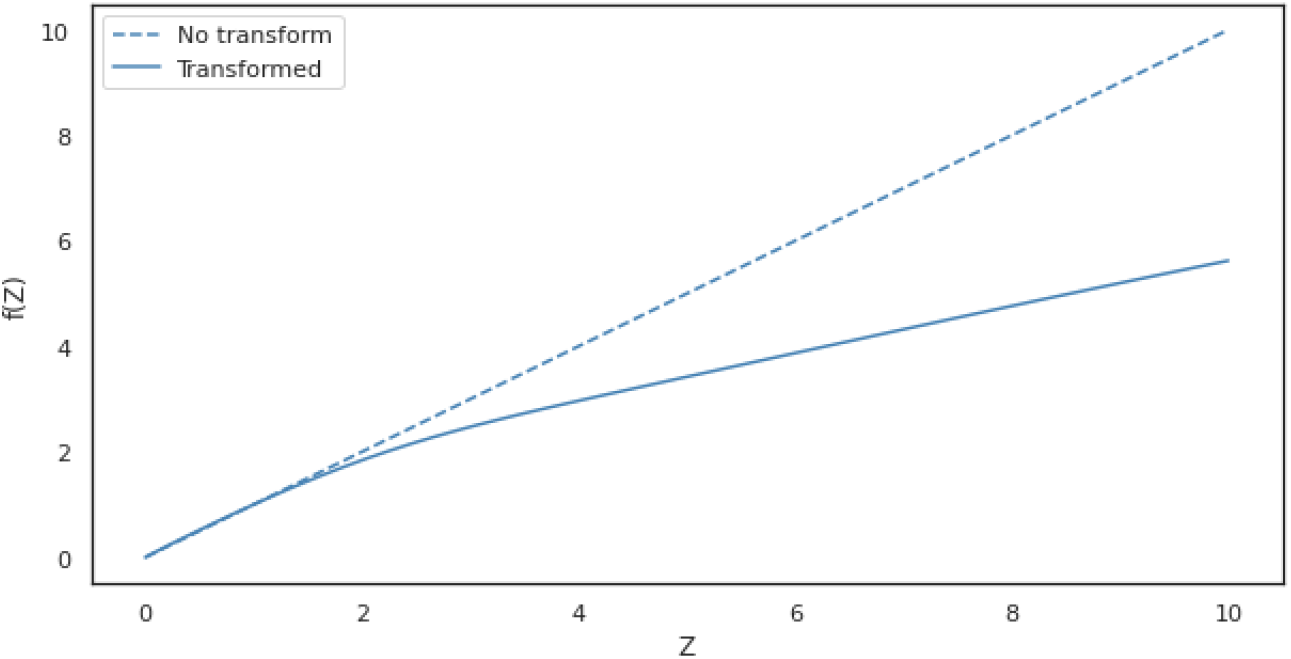
The effect of the transformation given by Equation 1 on the Z-scores.

### WES data

Whole Exome Sequencing (WES) data was acquired over a longer time period (2013-2021), and was performed either using the Agilent Clinical Research Exome V1 (sureselect SSCRE V1) or Agilent Clinical Research Exome V2 (sureselect SSCRE V2) on a *Illumina NovaSeq* sequencer using paired-end reads with a read-length of 150 bp. Reads were aligned to human reference genome build GRCh37/ hg19 (ucsc.hg19.nohap.fasta) using the BWA alignment algorithm (Li & Durbin, 2009). The VCF-files were obtained using GATK3 (McKenna, et al., 2010) and ANNOVAR was used to annotate gene names and variants (Wang, et al., 2010). All patients included in this study from which WES data was used gave consent for anonymous use of their data for research purposes.

### Retrieving human metabolic reactions

We used the KGML parser from https://github.com/biopython/biopython (20-03-2020) to process KEGG (Kanehisa, 2000) pathways and modules, where we filtered on reactions involved in humans (using the *hsa* pre-fix). When retrieving the KEGG networks, some reactions were associated with more than one enzyme, for which KEGG returns the same unique reaction as many times as it is associated with the different enzymes, leading to a multiplicity for these reactions. We removed this multiplicity but we remained all the associated enzymes with this reaction. In other words, in these cases the same S_R_ score for that reaction was assigned to all associated enzymes.

To increase the overlap between the metabolites measured in plasma and metabolites in in the pathways/modules (from KEGG), we manually added some reactions. These can be found in Supplement 4 Most of these reactions were obtained from Recon / Virtual Metabolic Human (Noronha, et al., 2018).

### Reafect

To determine the *deficient reaction score* for a certain reaction, we first consider the decay of the Z-score over a path *p* leading for metabolite *m* to reaction *R*:

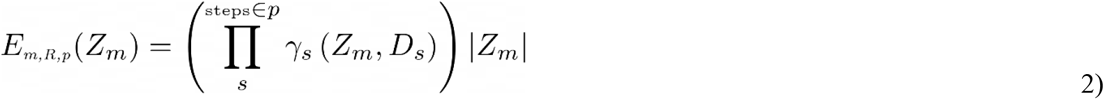

Here, *E*_*m,R,B*_ (*Z*_*m*_) is the *effective Z-score* for metabolite *m* from the perspective of reaction *R* along reaction path *p*. γ_*s*_ (*Z*_*m*_, *D*_*s*_) is the decay factor for step *s* and depends on the biochemical directionality of the step (*D*_*s*_) (upstream, downstream, reversible) and the sign of the Z-score (*Z*_*m*_):

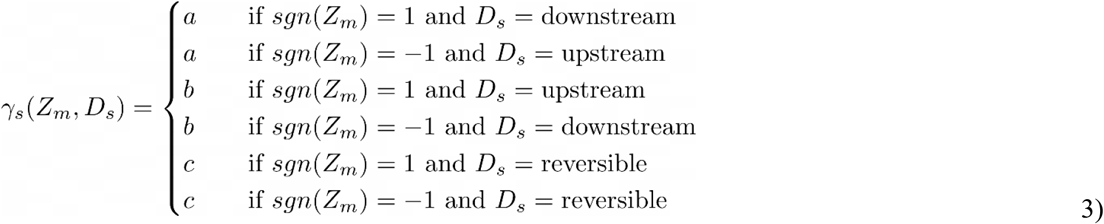

For irreversible reactions, *a* is the decay factor for the Z-score when the sign of the Z-score is positive and the reaction step is downstream, or when the Z-score is negative and the reaction step is upstream. The parameter *b* is the decay factor for the opposite cases; the sign of the Z-score is positive (negative) and the reaction step is upstream (downstream). For reversible reactions we introduce parameter *c* as decay factor. Note that this decay is independent of the sign of the Z-score.

Since more paths (*p’*s) could be possible between metabolite *m* and reaction *R*, and these could have different lengths, we calculated a normalized *effective Z-score* for every path:

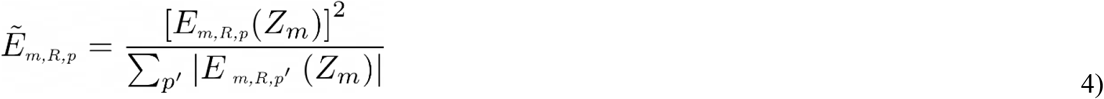

where 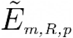 is the *normalized effective Z-score* for path *p*. The summation over *p*^***’***^ indicates all paths leading from *m* to *R*. In this way, paths originating from *m* with (relatively) low *effective Z-score* strengths (such as longer paths) are weighted less in the *normalized effective Z-score* whereas short paths get more weight since their *effective Z-score* is relatively large (when compared to the other paths). All paths (*p’*s) were determined by constructing an ‘ego graph’ around each metabolite, selecting a subset of neighboring metabolites and reactions around this central metabolite. To reduce computational cost, we set a limit of 15 reaction steps (metabolite-reaction or reaction-metabolite) around this ego graph, and a maximum of 10 paths for travelling from *m* to *R*.

Next, we summed all *normalized effective Z-scores* but we made a distinction between *normalized effective Z-scores* where its path is connected to the upstream or the downstream side of reaction *R*. For clarity, let us consider a direct substrate *m* of reaction *R*, which has a direct connection at the upstream side of the reaction. Let us also assume that there is a path going from *m*, via other reactions, which ends at the downstream side of the reaction. Since we have two paths, we have two *normalized effective Z-scores;* one belonging to the direct connection, the other belonging to the longer path. Since the latter path is longer, its *normalized effective Z-score* will be less than the *normalized effective Z-score* of the direct connection (Equation 4). We aggregated all metabolite *normalized effective Z-scores* based on the Z-score sign and connection to the reaction (downstream or upstream):

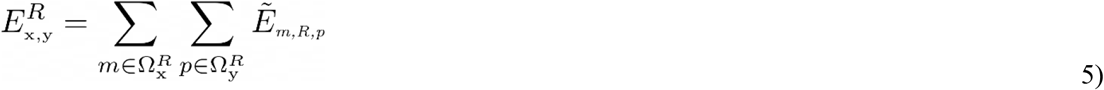

with, X ∈.{positive Z-score, negative Z-score}, *y* ∈ {downstream, upstream} 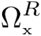 indicates the set of metabolites having a Z-score sign equal to *x* and 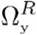 indicates the set of paths from *m* to *R* which are connected to the *y-*side of reaction *R* (downstream or upstream). Since reversible reactions lack a clear defined up – and downstream side, we assigned one of each side to the up – or downstream side while making sure that product/substrate information was conserved.

Finally, we defined the *deficient reaction score* for reaction as:

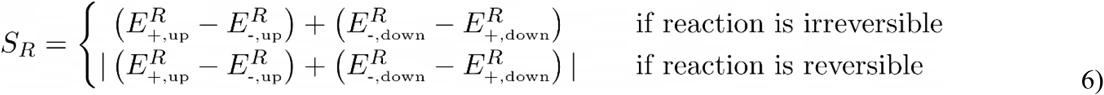

where we replaced ‘positive Z-score’ and ‘negative Z-score’ for the symbol ‘+’ and ’-’, respectively. We replaced ‘downstream’ and ‘upstream’ for ‘down’ and ‘up’, respectively. We observe that *S*_*R*_ increases for net positive *normalized effective Z-scores* located at the upstream side of the reaction and for net negative *normalized effective Z-scores* located at the downstream side of the reaction, while *S*_*R*_ decreases for the opposite cases. When a reaction is reversible we decided to take the absolute value, arguing that we are interested in an imbalance of the net positive and negative *normalized effective Z-scores* across the reaction regardless of which side of the reaction these *normalized effective Z-scores* were positioned.

We need to realize that some enzymes catalyze multiple (unique) reactions, which leads to a multiplicity of the S_R_ scores per enzyme and (potentially) shared reactions with other enzymes catalyzing the same reaction(s). In this study we dealt with this issue by taking the maximum occurring S_R_ score for each enzyme, even if that same S_R_ score was already assigned to another enzyme. Alternatively, we could have considered the use of another metric (other than the maximum) such as the average of all associated S_R_ scores, but since some associated reactions were considered poor, this average score could affect the performance negatively.

### Overall performance of Reafect using bootstrapped AUC

Annotation of metabolites in the metabolomics data was performed per batch, which resulted in an unequal number of annotations per batch. This difference also affected the number of unique enzymes on which ranking was based per patient (Figure 4, *Total number of enzymes*). To correct for this, we expressed the (absolute) rank in as a percentile by dividing by the total number of enzymes multiplied by 100%. The overall performance of *Reafect* for a certain choice of (*a, b, c*) was measured by displaying the percentage (vertical axis) of the IEM patients having the percentile rank of the correct IEM within the top x (horizontal axis). Calculating the area under the curve (AUC) for this relationship gives a measure for the overall performance, since a higher AUC indicates that a larger percentage of the IEM patients have a lower rank (steeper increase of the curve). We used a bootstrap procedure where we selected 1000 times a random 75% of the total IEM patients for which we calculated the AUC. By taking the 50^th^ percentile of these 1000 AUCs we obtained a more robust overall performance for each (*a, b, c*).

### MetPropagate and comparison with Reafect

We downloaded the weighted STRING network (v11) from https://github.com/emmagraham/metPropagate (07-08-2020). ME scores were calculated in the exact same manner as described by Linck et al. Using the same terminology, metabolites having |Z-score| > 1.5 where considered as ‘differentially abundant metabolites’. ME scores were propagated using the Local and Global Consistency (LGC) algorithm with settings `max_iter=30` and `alpha=0.99`.

To objectively compare *Reafect* with *MetPropagate* we took several factors into account:

1. Only metabolites were included with (HMDB) identifiers in the pathways/modules used by *Reafect* and which were also present in the gene-metabolite sets used by *MetPropagate*.
2. Before determining ranks, the propagated ME scores for every gene were assigned to the associated enzyme(s). We removed genes (and thus enzymes) which did not overlap in the output of both algorithms. Thus, both outputs contained the exact same number of unique enzymes on which ranking was performed.
3. The ranks for *MetPropagate* were calculated using the propagated ME scores on the enzyme level. Note that we took the maximum propagated ME score for an enzyme when more genes were associated with that enzyme. Similarly, the ranks for *Reafect* were determined from the S_R_ scores (as described above).

### CADD scores

Variants called by GATK3 (see Method, *WES data*) were annotated with *CADD* scores from Genome build GRCh37/ hg19 v1.6 (https://cadd.gs.washington.edu/download) for both SNVs and InDels. In this study we used the *CADD* (Phred) scores in two manners: 1) ranking genes based solely on the maximum *CADD* score occurring in each gene and 2) ranking genes using the *deficient reaction score* (S_R_ score) from *Reafect* combined with the *CADD* scores. Note, that only genes were included in this ranking for which a S_R_ score was determined and which were present in the WES data.

Gene ranking using *Reafect* in combination with *CADD* scores was done as follow:

1. Per enzyme the maximum S_R_ score was determined for all associated reactions. For each enzyme, all associated genes were determined and the same maximum S_R_ score was assigned to these genes.
2. The maximum *CADD* (Phred) score per gene was determined.
3. The S_R_ score (step 1) was multiplied with the *CADD* score (Phred) for each gene.
4. Genes were ranked on their integrated score (step 3).

For a subset of the IEM patients included in this study the disease-causing variant was identified either using whole exome sequencing (WES), Sanger sequencing or using an SNP array. Since WES data was not available for most IEM patients where the disease-causing variant(s) is identified, we assumed that we could include these patients using 15 random WES backgrounds while inserting the known disease-causing variant in each background. Consequently, we obtained 15 different rankings for each disease-causing gene. We assumed that the average of these 15 rankings is a good estimate of the rank when a real WES background was used (Discussion).

### Excluded IEM patients which were initially measured

Although some IEM patients were initially measured they were not included in this study, which had two main reasons. First, in some cases there was no (clear) associated reaction related to the metabolites known as biomarkers for that IEM, e.g. defects in cofactor metabolism. For example, we left out a patient with a mutation in the MMACHC gene, one with a mutation in the MOCS3 gene and two patients with glutaric acidemia type 2 (ETFDH, ETFA, ETFB). Secondly, since *Reafect* does not make a distinction between different compartments within the body or cell, the inclusion of enzymatic deficiencies related to transport proteins is complicated. In these transport reactions the metabolite itself does not change, only its location changes, and therefore build-up of these metabolites are expected only in certain parts of the body or cell (Discussion). For this reason, we were not able to include a few patients with lysinuric protein intolerance (SLC7A7), and a patient with organic cation transporter 2 deficiency (SLC22A5)

## Data Availability

Data is available on request. Software is available.

https://github.com/mbongaerts/Reafect

## Author contribution

The development of the methods was done by MB, MR and GR. RB performed all the experimental work, among which compound identification of the metabolomics data. MB developed the software and performed the computational experiments. WdV contributed in methods to analyze and process WES data. The interpretation of the results was done by MB, MR, HB and GR. The manuscript was written by MB, MR, HB and GR. SD, JL, HH and MW provided data and resources. The research was under supervision of GR.

## Acknowledgement

We want to thank and acknowledge Professor Robert Hofstra for his support and Dr. Geert Geeven for his comments and feedback on the manuscript.

## Code availability

*Reafect* is available at https://github.com/mbongaerts/Reafect

## Supplementary data

### Supplement 1. Comparison of ranks IEM patients *Reafect* versus *MetPropagate*

**Figure S1.**
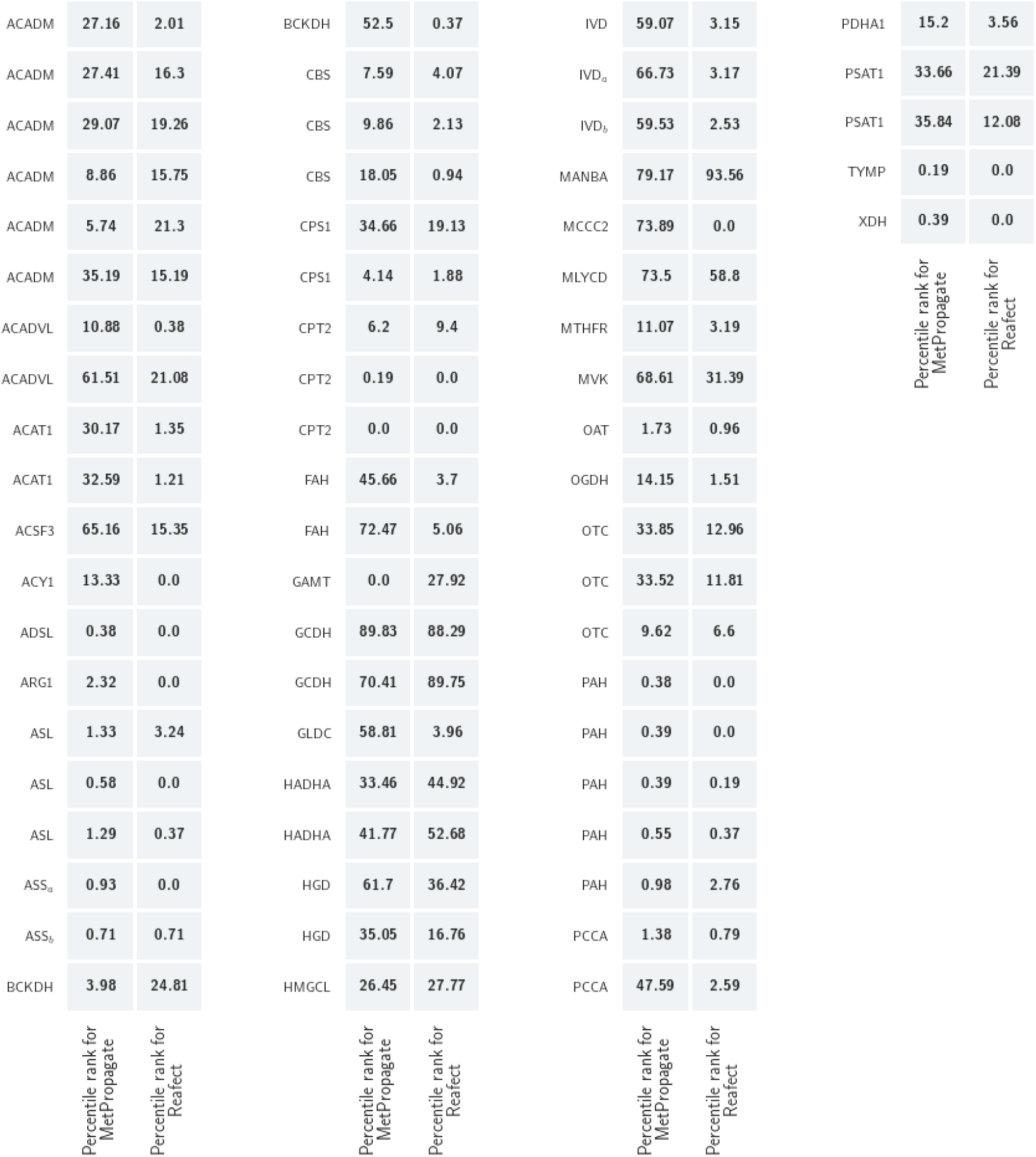
Comparison of the percentile ranks between *Reafect* and *MetPropagate* per IEM patient.

### Supplement 2. The effect of flipping the Z-score on IEM ranking performance

To explore the importance of taking the biochemical directionality and the sign of the Z-scores into account, we flipped the sign for all Z-scores (in all patients), and used *Reafect* to rank the enzymatic deficiencies (Figure S2). We observe that the AUC was reduced by 26%, and for the lower region of the performance curve (<10%), the partial AUC even dropped by 61%. These results underline the importance of including this information when considering IEM ranking algorithms. A detailed comparison between the ranks obtained from both approaches can be found in Figure S3.

**Figure S2.**
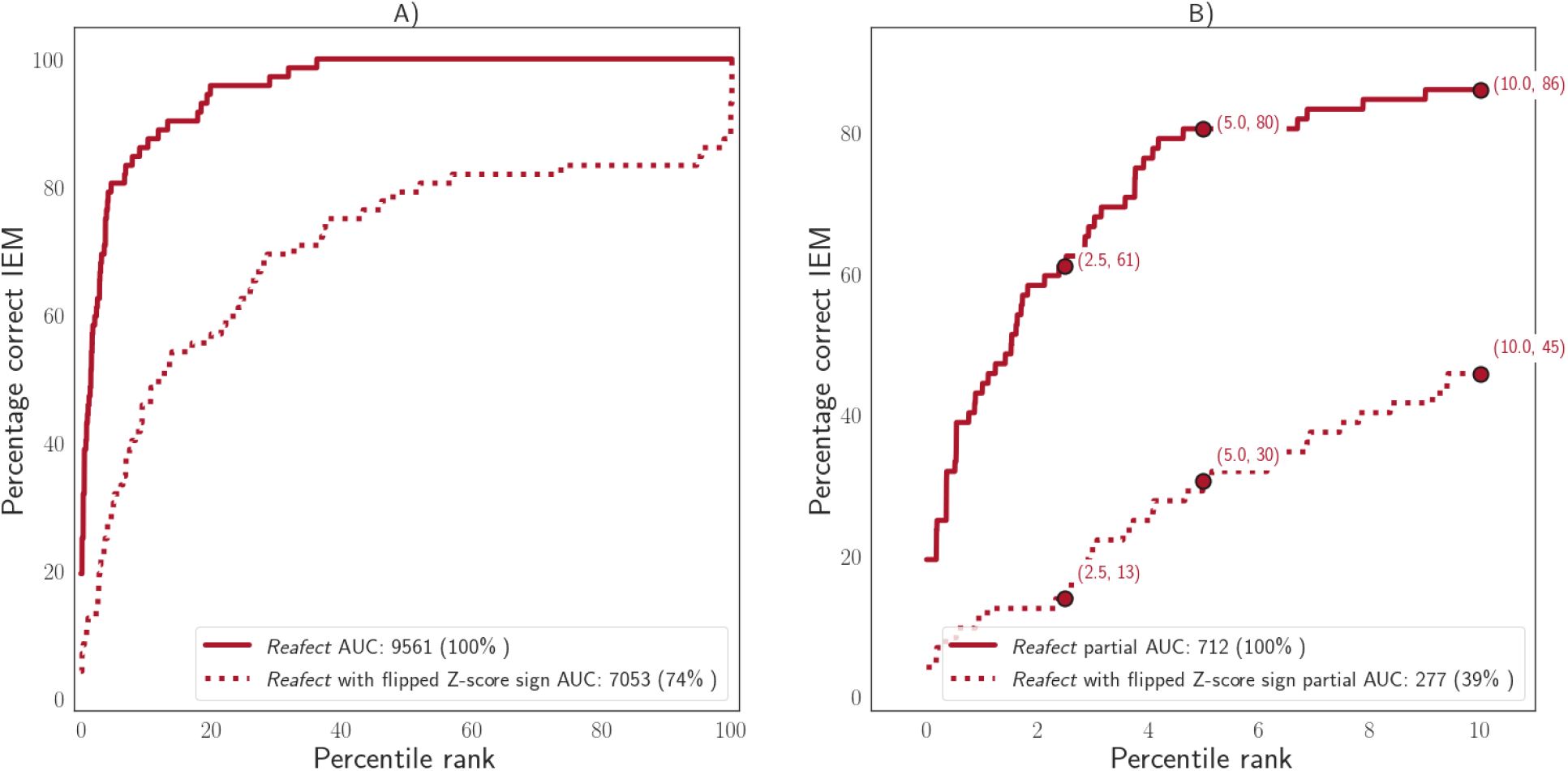
A) Full performance curves for *Reafect* and *Reafect with flipped Z-score signs*. B) Percentile ranks <= 10%.

**Figure S3.**
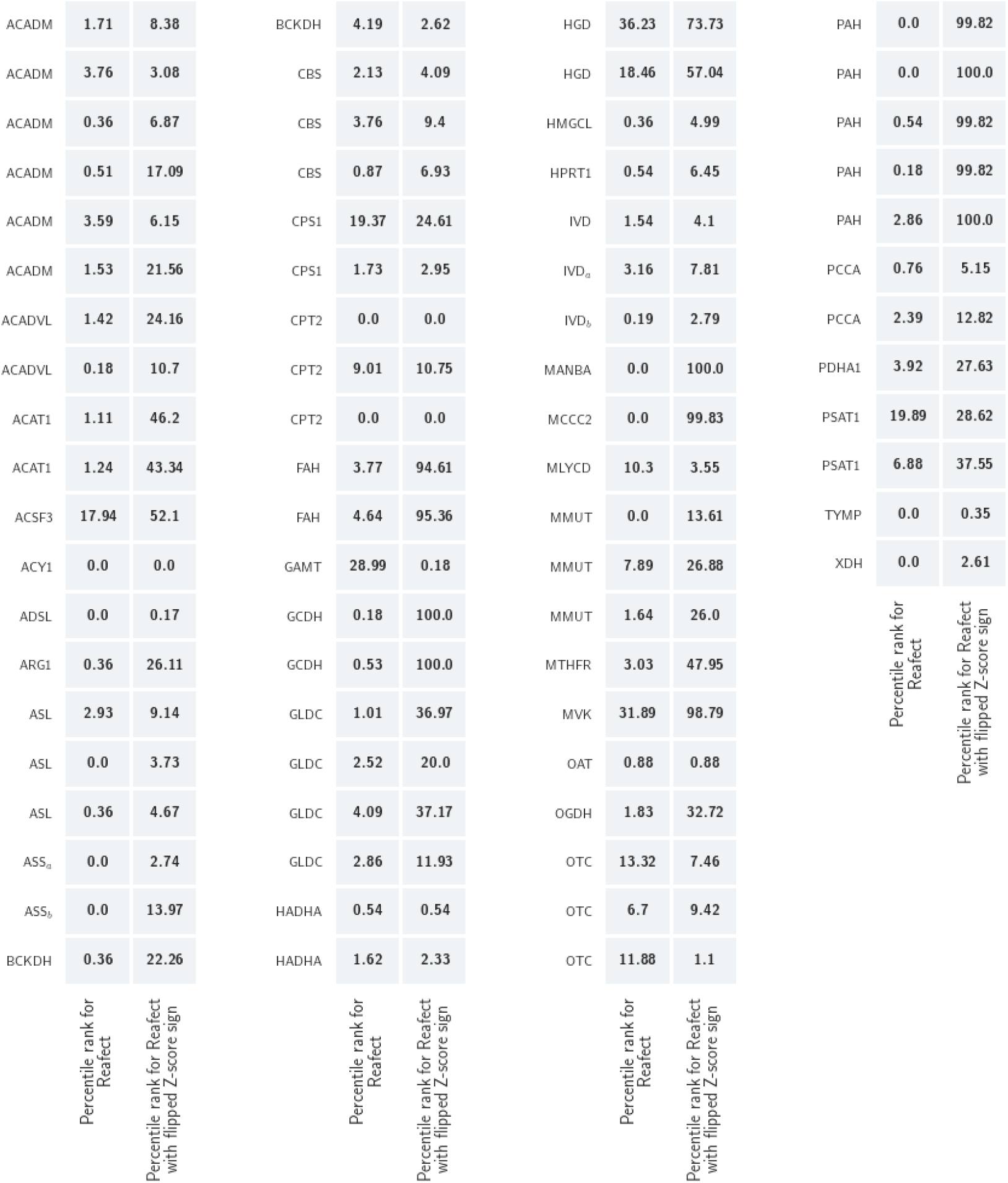
Comparison of the percentile ranks between *Reafect* and *Reafect with flipped Z-score signs* per IEM patient. We clearly observe the negative effect of reversing the sign of the Z-scores on ranking.

### Supplement 3. *CADD* scores for WES and pathogenic variants

**Figure S4.**
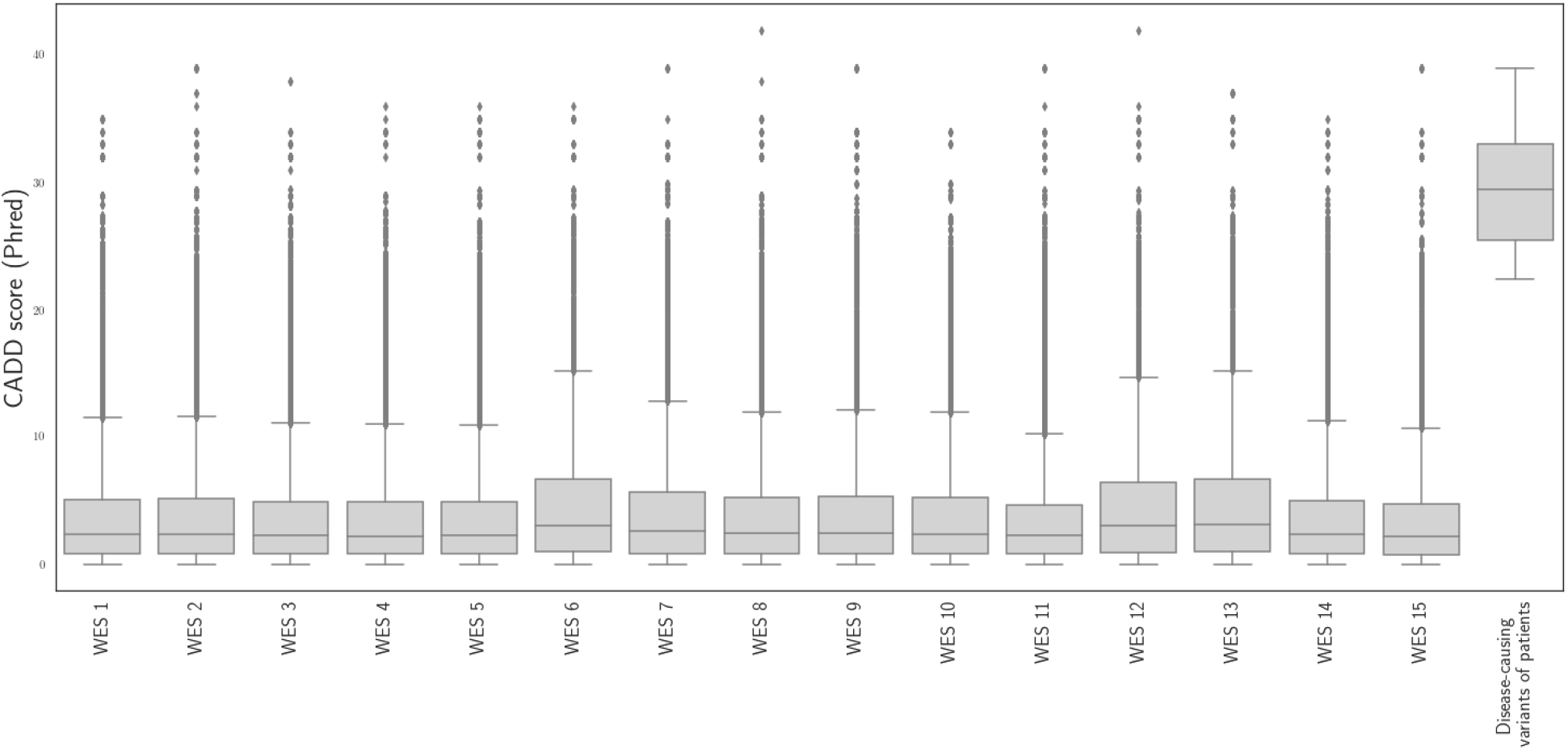
Each boxplot indicates the distribution of the *CADD* (Phred) scores for variants in metabolic genes obtained in 15 random WES files. The last boxplot shows the *CADD* scores (Phred) for the disease-causing variants found in the IEM patients (Table 1).

### Supplement 4. Manually added reactions

The KEGG pathways and modules were extended with some additional reactions (see Table S1) to increase the overlap between metabolites present in the pathways/modules and metabolites measured in plasma. Note, that a reaction is defined as a graph which also includes a reaction node.

**Table S1.** Manually added reactions. The second and fourth column indicate the directionality of the reaction. ‘<=>’ indicates that the reaction is reversible whereas ‘=>’ indicates the direction of an irreversible reaction. Note that these ‘reactions’ passes through a reaction node (Reaction ID). Most reactions originate from Recon3D.

| Metabolite 1 |  | Reaction ID |  | Metabolite 2 |
| --- | --- | --- | --- | --- |
| 3-Hydroxypyruvic acid | => | HPYRR2x | => | Glyceric acid |
| 2-Methylbutyrylglycine | <=> | RE2428M | <=> | 2-Methylbutanoyl-CoA |
| 2-Methylbutyrylglycine | <=> | RE2428M | <=> | Glycine |
| C16OH \| 3-Hydroxyhexadecanoylcarnitine | <=> | C16OHc | <=> | (S)-3-Hydroxyhexadecanoyl-CoA |
| 3-Methylcrotonylglycine | <=> | RE2111M | <=> | 3-Methylcrotonyl-CoA |
| 3-Methylcrotonylglycine | <=> | RE2111M | <=> | Glycine |
| glnac-man | => | B_MANNASEly | => | N-Acetyl-D-glucosamine |
| glnac-man | => | B_MANNASEly | => | D-Mannose |
| C10 \| Decanoylcarnitine | <=> | C10CPT1 | <=> | Decanoyl-CoA |
| Glycylproline | <=> | GLYPROPRO1c | <=> | Glycine |
| Glycylproline | <=> | GLYPROPRO1c | <=> | Proline |
| C6 \| Hexanoylcarnitine | <=> | C6CPT1 | <=> | Hexanoyl-CoA |
| Homocysteine thiolactone | <=> | RE1933C | <=> | Homocysteine |
| Isobutyrylglycine | <=> | RE2429M | <=> | Glycine |
| Isobutyrylglycine | <=> | RE2429M | <=> | 2-Methylpropanoyl-CoA |
| C5 \| Isovalerylcarnitine | <=> | C5CPT1 | <=> | 3-Methylbutanoyl-CoA |
| Isovalerylglycine | <=> | RE2427M | <=> | Glycine |
| Isovalerylglycine | <=> | RE2427M | <=> | 3-Methylbutanoyl-CoA |
| Malonyl-CoA | => | r0430 | => | C3DC \| Malonylcarnitine |
| N-Acetylaspargine | <=> | RE2032M | <=> | Asparagine |
| C14 \| Tetradecanoylcarnitine | <=> | C14CPT1 | <=> | Tetradecanoyl-CoA |
| C5:1 \| Tiglylcarnitine | <=> | C51CPT1 | <=> | 2-Methylbut-2-enoyl-CoA |
| Octanoyl-CoA | <=> | C8CPT1 | <=> | C8 \| Octanoylcarnitine |
| Butanoyl-CoA | <=> | C4CPT1 | <=> | C4 \| Butyrylcarnitine |
| Propanoyl-CoA | <=> | C3CPT1 | <=> | C3 \| Propionylcarnitine |
| (2S,3S)-3-Hydroxy-2-methylbutanoyl-CoA | <=> | R_2M3HBUC | <=> | 2-Methyl-3-hydroxybutyric acid |
| 2-Methylbut-2-enoyl-CoA | <=> | R_TIGGLYc | <=> | Tiglylglycine |
| N-Acetylmethionine | <=> | RE2640C | <=> | Methionine |
| N-Acetylalanine | <=> | RE2642C | <=> | L-Alanine |
| Glutaryl-CoA | <=> | FAOXC5C5DCc | <=> | C5DC \| Glutaryl carnitine |
| 3-Methylglutaconyl-CoA | <=> | 3mgcoac61dcmgcrn | <=> | C6:1DC \| 3-Methylglutaconylcarnitine |
| 3-Methylglutaconyl-CoA | <=> | MGCHrm | <=> | (S)-3-Hydroxy-3-methylglutaryl-CoA |
| (S)-3-Hydroxy-3-methylglutaryl-CoA | <=> | hmgcoac6dcmgcrn | <=> | C6DC \| 3-Methylglutaryl carnitine |
| 3-Hydroxyisovaleryl-CoA | <=> | C059983ivcrn | <=> | C5OH \| 3-Hydroxyisovalerylcarnitine |
| 3-Hydroxyisovaleryl-CoA | <=> | C059983CE2028 | <=> | 3-Hydroxyisovaleric acid |
| Adenylosuccinate | => | C03794succinyladenosine | => | Succinyladenosine |
| L-Aspartate | => | ASPCTr | => | N-Carbamoyl-L-aspartate |
| Carbamoylphosphate | => | ASPCTr | => | N-Carbamoyl-L-aspartate |
| Dihydroorotic acid | => | DHORTS | => | N-Carbamoyl-L-aspartate |
| Dihydroorotic acid | <=> | DHORD9 | <=> | Orotic acid |
| Malonate | => | C00383malcoa | => | Malonyl-CoA |
| Malonyl-CoA | => | MCDm | => | Acetyl-CoA |
| 7-Dehydrocholesterol | => | HMR_2114 | => | Vitamine D3 |
| Cholesterol sulfate | <=> | RE1100L | <=> | Cholesterol |

### Supplement 5. Optimizing the decay factors while excluding the samples with an identified disease-causing variant

In this study we used all 72 IEM patients samples to optimize the decay factors and to evaluate *Reafect*’s performance on IEM/gene ranking. To support our findings, especially for the results where we integrated the *deficient reaction scores* with *CADD* scores (Table 1), we optimized the decay factors on 44/72 samples where we excluded the patients with an identified disease-causing variant. We used the same bootstrap procedure for determining the optimal values for the decay factors (Methods). We found that the optimum was at *a* = 0.85, *b* = 0.45, *c* = 0.75 (100%), and the second best combination was *a* = 0.85, *b* = 0.35, *c* = 0.75 (99.96%) (Figure S5). However, we also observe that the optimum is wider and less well-defined as the one observed in Figure 2.

**Figure S5.**
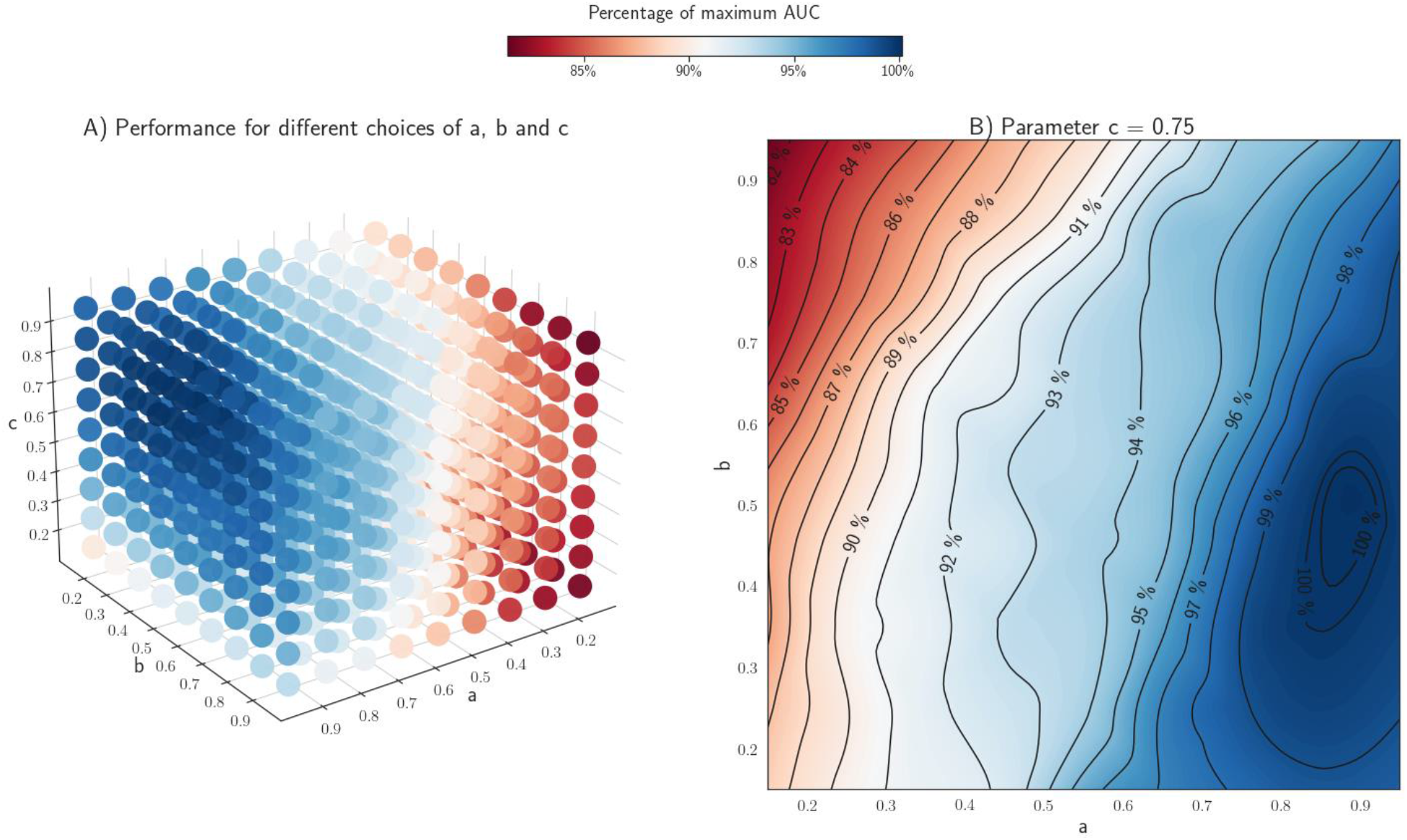
**A)** Bootstrapped AUCs (Methods) for given combinations of (*a,b,c*) indicating the performance of *Reafect*. The colors indicate the percentage of the maximum obtained AUC. For this analysis we used 44/72 IEM patients samples, where we excluded the patients with an identified disease-causing variant. **B)** Contour plot of the (cubic interpolated) bootstrapped AUCs while fixing *c=0*.*75* and varying *a* and *b*. The contour levels indicate the percentage of the maximum AUC reached at *a = 0*.*85, b = 0*.*45, c = 0*.*75*.

### Supplement 6. Contribution of subtle metabolite Z-scores on IEM ranking

We explored the contribution of more subtle metabolite Z-scores to the IEM ranking performance of *Reafect*. This was investigated by creating performance curves for various Z-score cutoffs, where we included only metabolite Z-scores for which |Z-score| < cutoff (Figure S6A) or |Z-score| > cutoff (Figure S6B). These results show that for decreasing cutoff values and |Z-score| < cutoff, the overall performance on IEM ranking also declines. This can be understood by realizing that for decreasing cutoff values, also more informative (disease-related) metabolites are excluded. More importantly, we observe that even for the lower cutoff values the overall performance is still positive (above the diagonal line), suggesting that more subtle metabolite Z-scores also contribute to IEM ranking. The same conclusion can be drawn from the experiment where we included only metabolites having a |Z-score| > cutoff. When increasing the cutoff values, we observe that the IEM ranking performance also decreases. Since in these cases only more extreme Z-scores are available for ranking, we conclude that more subtle metabolite Z-scores normally also contribute to IEM ranking.

**Figure S6.**
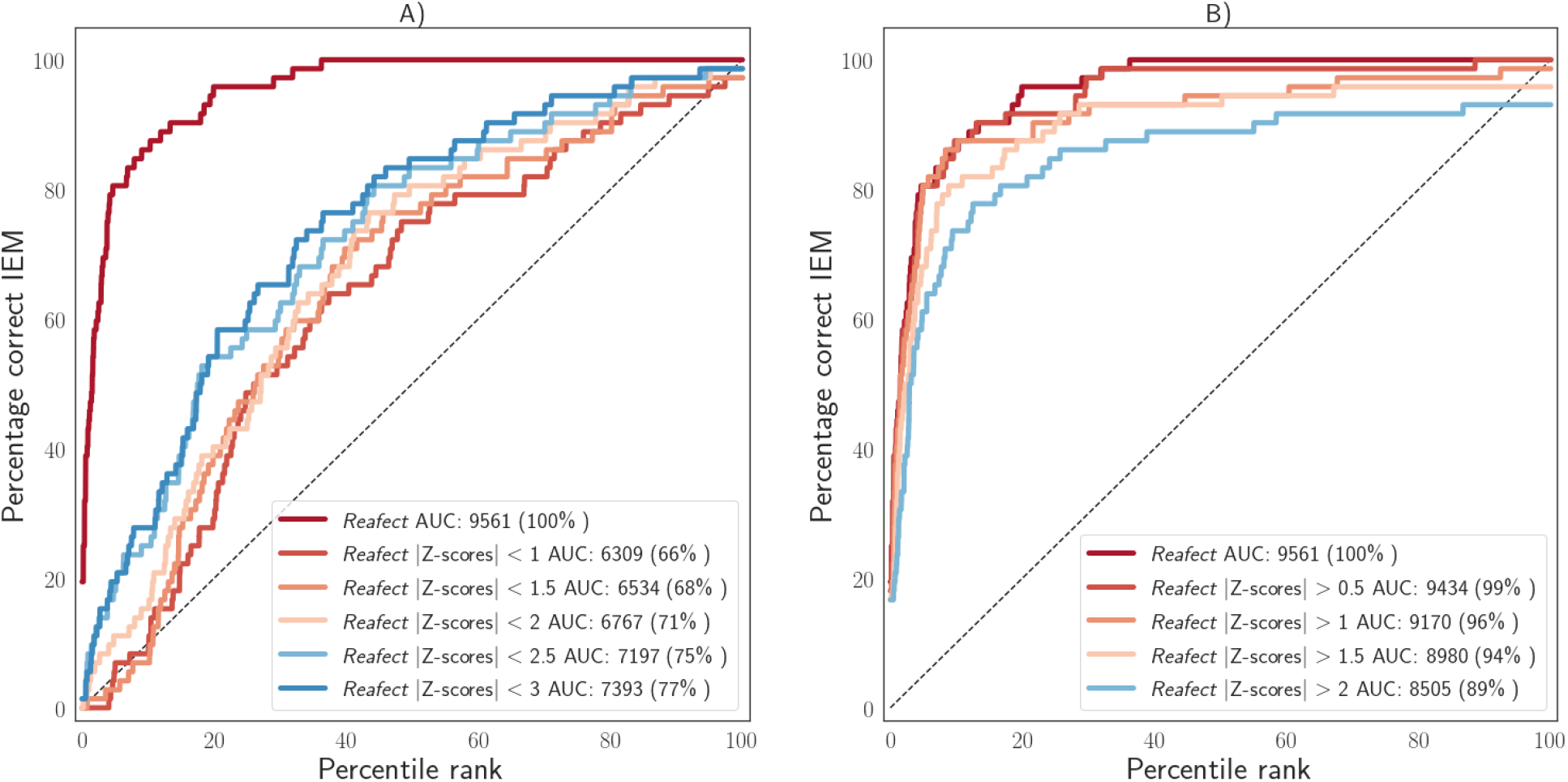
**A)** Full performance curves for *Reafect* for various Z-score cutoff values, and |Z-score| < cutoff. Cutoff values are indicated by the legend. **B)** Full performance curves for *Reafect* for various Z-score cutoff values, and |Z-score| > cutoff.

## References

Alaimo, J. T. et al., 2020. Integrated analysis of metabolomic profiling and exome data supplements sequence variant interpretation, classification, and diagnosis. Genetics in Medicine, 5.

Baumgartner, C. et al., 2004. Supervised machine learning techniques for the classification of metabolic disorders in newborns. Bioinformatics, 6, Volume 20, p. 2985–2996.

Bongaerts, M. et al., 2020. Using Out-of-Batch Reference Populations to Improve Untargeted Metabolomics for Screening Inborn Errors of Metabolism. Metabolites, 12, Volume 11, p. 8.

Bonte, R. et al., 2019. Untargeted Metabolomics-Based Screening Method for Inborn Errors of Metabolism using Semi-Automatic Sample Preparation with an UHPLC-Orbitrap-MS Platform. Metabolites, 11, Volume 9, p. 289.

Haijes, H. A. et al., 2020. Untargeted Metabolomics for Metabolic Diagnostic Screening with Automated Data Interpretation Using a Knowledge-Based Algorithm. International Journal of Molecular Sciences, 2, Volume 21, p. 979.

Kanehisa, M., 2000. KEGG: Kyoto Encyclopedia of Genes and Genomes. Nucleic Acids Research, 1, Volume 28, p. 27–30.

Kerkhofs, M. H. P. M. et al., 2020. Cross-Omics: Integrating Genomics with Metabolomics in Clinical Diagnostics. Metabolites, 5, Volume 10, p. 206.

Lee, J. J. Y. et al., 2017. Knowledge base and mini-expert platform for the diagnosis of inborn errors of metabolism. Genetics in Medicine, 7, Volume 20, p. 151–158.

Li, H. & Durbin, R., 2009. Fast and accurate short read alignment with Burrows-Wheeler transform. Bioinformatics, 5, Volume 25, p. 1754–1760.

Linck, E. J. G. et al., 2020. metPropagate: network-guided propagation of metabolomic information for prioritization of metabolic disease genes. npj Genomic Medicine, 7.Volume 5.

McKenna, A. et al., 2010. The Genome Analysis Toolkit: A MapReduce framework for analyzing next-generation DNA sequencing data. Genome Research, 7, Volume 20, p. 1297–1303.

Messa, G. M. et al., 2020. A Siamese neural network model for the prioritization of metabolic disorders by integrating real and simulated data. Bioinformatics, 12, Volume 36, p. i787–i794.

Noronha, A. et al., 2018. The Virtual Metabolic Human database: integrating human and gut microbiome metabolism with nutrition and disease. Nucleic Acids Research, 10, Volume 47, p. D614–D624.

Pirhaji, L. et al., 2016. Revealing disease-associated pathways by network integration of untargeted metabolomics. Nature Methods, 8, Volume 13, p. 770–776.

Pronicka, E. et al., 2016. New perspective in diagnostics of mitochondrial disorders: two years’ experience with whole-exome sequencing at a national paediatric centre. Journal of Translational Medicine, 6.Volume 14.

Rentzsch, P. et al., 2018. CADD: predicting the deleteriousness of variants throughout the human genome. Nucleic Acids Research, 10, Volume 47, p. D886–D894.

Stavropoulos, D. J. et al., 2016. Whole-genome sequencing expands diagnostic utility and improves clinical management in paediatric medicine. npj Genomic Medicine, 1.Volume 1.

Thiele, I. et al., 2013. A community-driven global reconstruction of human metabolism. Nature Biotechnology, 3, Volume 31, p. 419–425.

Wang, K., Li, M. & Hakonarson, H., 2010. ANNOVAR: functional annotation of genetic variants from high-throughput sequencing data. Nucleic Acids Research, 7, Volume 38, p. e164–e164.

Waters, D. et al., 2018. Global birth prevalence and mortality from inborn errors of metabolism: a systematic analysis of the evidence. Journal of Global Health, 11.Volume 8.

Wright, C. F., FitzPatrick, D. R. & Firth, H. V., 2018. Paediatric genomics: diagnosing rare disease in children. Nature Reviews Genetics, 2, Volume 19, p. 253–268.

